# Evaluating methods to quantify the effect of deprescribing a medication on patient clinical outcomes using a simulation study

**DOI:** 10.64898/2026.11.02.26367054

**Authors:** Ryan Muddiman, Teresa Pérez Pérez, Florencia Inés Aiello Battan, Emma Wallace, Tom Fahey, Mary E. Walsh, Fiona Boland, Li Wei, Frank Moriarty

## Abstract

Observational studies using electronic health records are being increasingly used to answer research questions of interest in deprescribing, where a medication is intentionally discontinued or the dose reduced. Target trial emulation (TTE) is particularly attractive as it is guided by sound theoretical principles based on causal inference. In the field of deprescribing, the evaluation of TTE within a simulation setting can provide evidence on optimal approaches for implementation and compare different estimation approaches which theory cannot easily inform. Deprescribing studies are susceptible to time-dependent confounding, competing risks due to early adverse events, time-dependent effects and other data complexities which may render estimates biased if such aspects are not properly addressed. In this work, a Monte-Carlo simulation study was conducted to estimate the performance (bias and coverage) of seven methods within several synthetic data generating mechanisms involving the above characteristics. The methods implemented were Cox regression, inverse probability of treatment weighted (IPTW) Cox regression, naive sequential Cox regression, IPTW weighted sequential Cox regression, extended Cox regression and the parametric g-formula. The IPTW Cox regression and IPTW sequential Cox regression were the only methods achieving nominal coverage and unbiasedness under the scenarios evaluated.

## 1 INTRODUCTION

Deprescribing is defined as stopping or tapering a medication that is considered by a medical practitioner to be of no clinical benefit. ^1^ As it is the removal of a therapy, there are no regulatory requirements for demonstrating the efficacy or safety of deprescribing using, for example, conclusive randomised controlled trials (RCTs). Several RCTs have been conducted in order to determine the risks and benefits of deprescribing certain drug classes compared to continuation. While RCTs are the gold standard for assessing the causal effect of an intervention, they are particularly difficult to conduct when examining the deprescribing of medicines, since the eligible population of interest are typically older with significant multi-morbidity; hence are inherently more vulnerable than younger patients.

To address these challenges, observational data such as electronic health records (EHR) may be analysed in an attempt to estimate causal effects. ^2^ In non-randomised studies, the risk of bias is inherently greater than in a randomised controlled trial. Residual confounding between experimental and comparison groups, as well as heterogeneous treatment effects may lead to incorrect conclusions depending on modelling choices. In deprescribing interventions specifically, there is often a highly unpredictable treatment course, due to the lack of clinical guidance, and intercurrent events may occur such as restarting drug treatment, that may modify the probability and/or interpretation of the outcomes of interest. Such events must be accounted for in the causal question to prevent biased inference.

When studying the effect of drug discontinuation a new-user design may be used, where the control group is those continuing on the medication of interest and the exposed group are those for whom deprescribing of the medication is initiated. However, deprescribing may not be initiated for certain people such as those who do not experience major adverse events, and so determining the effects in these patients may be difficult without interventional data. Within deprescribing studies, the outcomes of interest may range from the occurrence of major adverse events (e.g., acute myocardial infarction, stroke, cardiovascular death) to indicators of medication burden (e.g., drugs with significant anticholinergic or sedative effects). The cause-effect relationship between interventions and clinical outcomes is typically summarised using time-to-event methods, incorporating not only whether the event occurs but also when it occurs.

From a time-to-event analysis perspective, there are several quantitative approaches for estimation of causal effects in nonrandomised settings, and the G-methods are necessary for valid inference when treatment affects the failure time of interest. ^3^ The application of G-methods in observational studies on deprescribing is natural due to the time-dependent nature of treatment, where time-dependent confounding may be present. However, to date, the performance of the various methods under realistic data scenarios has not been tested.

Target trial emulation (TTE) is being increasingly recognised as a framework which may enable the estimation of causal effects, given certain assumptions, similar to those that may be obtained from an RCT. ^4^ In TTE, a question-first approach is taken by specifying the causal question, defining inclusion criteria and treatment strategies for a hypothetical RCT which would successfully generate the data required to answer the question, and then determining the modelling/data requirements. While TTE is not a quantitative method of estimation, it can be considered a study design or framework which formalizes the notion of the causal estimand and should impact the definition of risk sets as well as estimation in the analysis stage.

The aim of this study is to evaluate the performance of several estimation methods within the TTE framework for causal inference using semi-synthetic data. The simulated data represents longitudinal event histories which exhibit time-dependent confounding, stochastic treatment switching and terminal competing risks. We define the data generating mechanism in section 5 using a conceptual model of how event histories may be generated in routine care. We then describe the details of a target trial based on a clinical research question that may be emulated using the observed data presented in section 6. The research question focusses on the effect of deprescribing versus continuing a medication, on the occurrence of a major adverse event

## 2 THE TARGET TRIAL

A hypothetical pragmatic target trial focused on aspirin deprescribing was specified using the TARGET (Transparent Reporting of Observational Studies Emulating a Target Trial) framework ^5^ and is presented in Table 1. TARGET recommends specifying the eligibility criteria, treatment strategy, assignment, follow-up, outcomes, causal contrasts, identifying assumptions and data analysis plan. While not all of these attributes affect estimation or are relevant to the simulation, we give an example of a full target trial specification for context. Within healthcare research, estimands are the target of inference and are composed of five components ^6^; Population (defined by eligibility criteria), intervention or treatment conditions (treatment strategies), endpoint (outcomes), summary measure and strategies for handling intercurrent events. Intercurrent events are those post-treatment events which may modify the chance of occurrence or interpretation of the measurements related to the outcomes of interest. All estimand components are specified in Table 1. The number of estimands in a TTE specification corresponds to the number of distinct targets of inference, and in this study, two estimands were defined.

**TABLE 1.**
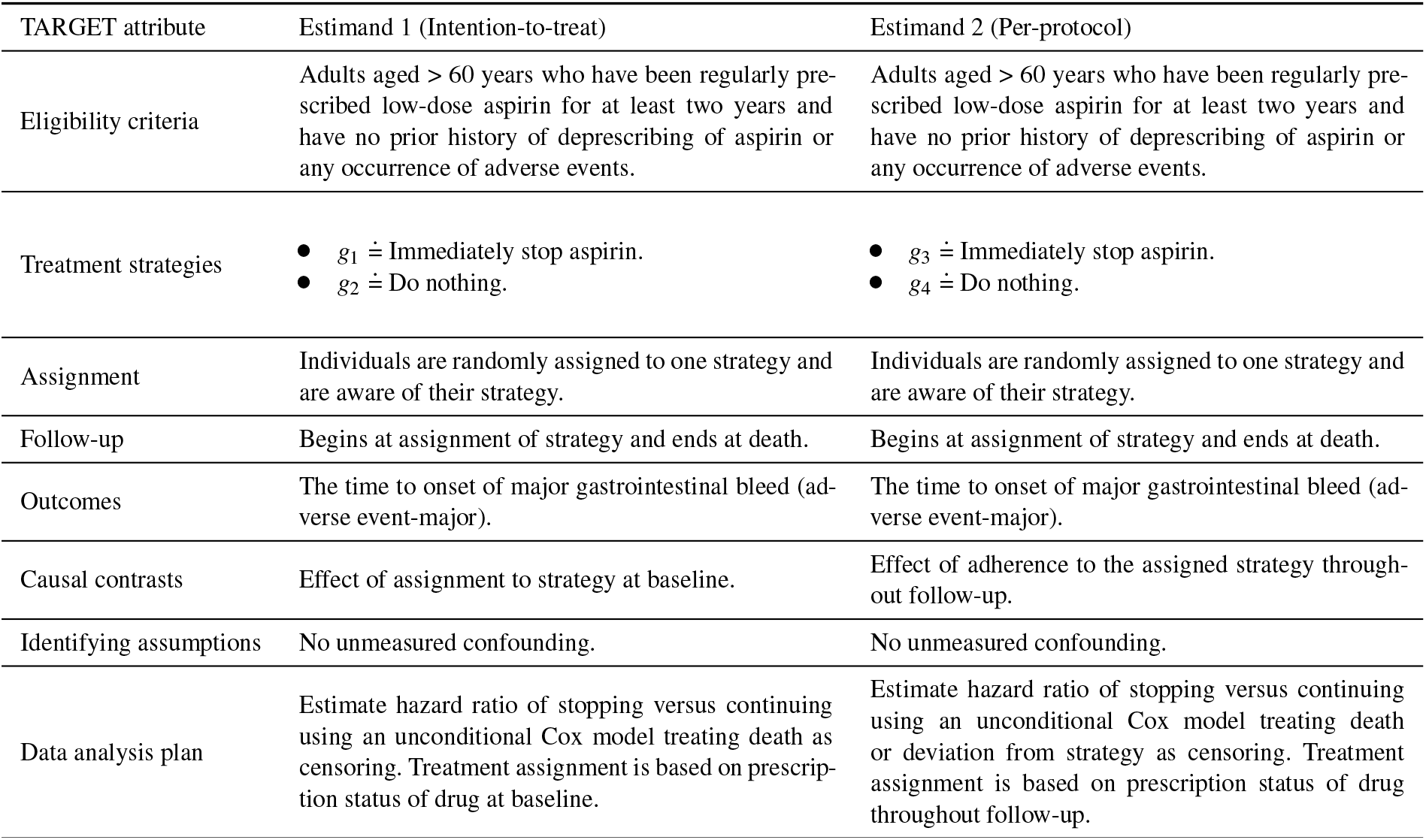
Specification of a hypothetical target trial for a research question focusing on deprescribing effectiveness for long-term aspirin users.

| TARGET attribute | Estimand 1 (Intention-to-treat) | Estimand 2 (Per-protocol) |
|---|---|---|
| Eligibility criteria | Adults aged > 60 years who have been regularly pre-scribed low-dose aspirin for at least two years and have no prior history of deprescribing of aspirin or any occurrence of adverse events. | Adults aged > 60 years who have been regularly pre-scribed low-dose aspirin for at least two years and have no prior history of deprescribing of aspirin or any occurrence of adverse events. |
| Treatment strategies | • *g*<sub>1</sub> ≐ Immediately stop aspirin.<br>• *g*<sub>2</sub> ≐ Do nothing. | • *g*<sub>3</sub> ≐ Immediately stop aspirin.<br>• *g*<sub>4</sub> ≐ Do nothing. |
| Assignment | Individuals are randomly assigned to one strategy and are aware of their strategy. | Individuals are randomly assigned to one strategy and are aware of their strategy. |
| Follow-up | Begins at assignment of strategy and ends at death. | Begins at assignment of strategy and ends at death. |
| Outcomes | The time to onset of major gastrointestinal bleed (ad-verse event-major). | The time to onset of major gastrointestinal bleed (ad-verse event-major). |
| Causal contrasts | Effect of assignment to strategy at baseline. | Effect of adherence to the assigned strategy through-out follow-up. |
| Identifying assumptions | No unmeasured confounding. | No unmeasured confounding. |
| Data analysis plan | Estimate hazard ratio of stopping versus continuing using an unconditional Cox model treating death as censoring. Treatment assignment is based on prescrip-tion status of drug at baseline. | Estimate hazard ratio of stopping versus continuing using an unconditional Cox model treating death or deviation from strategy as censoring. Treatment assignment is based on prescription status of drug throughout follow-up. |

Estimand 1 is typically called the intention-to-treat effect in the literature, as it quantifies the effect of initiation of treatment at a single point in time. Estimand 2 is typically called the per-protocol effect, since it quantifies the effect of following a specified treatment over time. Defining estimands using this nomenclature has been superseded by the estimands framework, which specifically requires defining how intercurrent events are handled. Intercurrent event handling is the only difference between estimand 1 and estimand 2 above. The TARGET statement does not specifically mention intercurrent events, but rather only mentions competing events. However, some intercurrent events are not competing events, for example use of additional medication may render the later measurements of the variable irrelevant or difficult to interpret but do not preclude the existence of the outcome. ^7^ As given in the follow-up section of Table 1, both estimands implicitly incorporate the so-called while-on-treatment strategy to deal with the competing event of death. Estimand 1 also uses the treatment policy strategy to ignore treatment switching which may occur after baseline i.e., restarting a drug due to adverse events. The treatment strategies in Table 1 are realistic and common strategies evaluated in deprescribing RCTs. A systematic review of drug discontinuation methodologies found that 28% of studies examined complete drug cessation as the intervention ^8^. In observational studies specifically, 84% examined abrupt discontinuation ^9^, highlighting the predominance of abrupt stopping in the existing evidence base. All four strategies are deterministic static strategies, since they do not depend on patient information (e.g., age) or random variables. Estimand 2 also incorporates sustained strategies, since continued treatment after baseline is part of the causal contrast.

## 3 CAUSAL INFERENCE IN NON-MARKOV MULTI-STATE MODELS

Multi-state modelling is a useful starting point for analysing and generating complex event histories such as those that may be found in observational deprescribing data. In this work, the data generating mechanism (DGM) is based on a multi-state model and we also incorporate such models in one of the estimation methods. Multi-state models are highly flexible models capable of describing processes that may change over time, such as an individual’s health status. A multi-state model is a generalization of the competing risks model, which generalizes the basic survival model. ^10^. Thus, multi-state modelling is a useful starting point for analysing and generating complex event histories such as those that may be found in observational deprescribing data. While popular for prediction, these models are not typically used for causal inference. ^11^ However, certain parameters of multi-state models can be given a causal interpretation when the assumptions of causal inference hold. Additionally, central to the utility of such models is a property known as modularity or local independence, which means that interventions on a causal node (e.g., modification of a transition intensity) does not directly affect other transition intensities. ^12,13^ This assumption is required to hold in order for the effects of interventions on transition intensities to be identifiable. Multi-state modelling approaches have been used to describe randomised controlled trials, ^14,15,16^ however, to our knowledge, they have not been used within the target trial emulation framework for conducting observational analyses.

A multi-state regression model can incorporate an arbitrary number of states, but as the number of states becomes larger it may become computationally infeasible to use them. The number of states is usually kept as small as possible, with the potential for extension including covariates that may modify the behaviour of transitions. Irreversible transitions, where a state can only be entered once are common (e.g., the three-state healthy-diseased-dead model), while reversible models, which are suited to e.g., relapsing-remitting conditions are less popular. A notable characteristic of reversible models is that the maximum number of state transitions is theoretically unbounded, which requires consideration for inference. ^17^

The average treatment effect (ATE) is a causal quantity and within a multi-state model it can be generally defined as any contrast of expectations of a functional of the stochastic process. For example, when the difference scale is of interest,

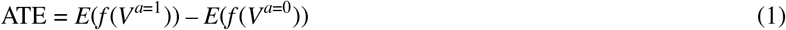

where *f* (*V*) may be a marginal process parameter such as restricted mean time spent in state *V* or a conditional parameter such as the hazard function of the time to first entry to a particular state, when interpreted functionally, i.e. comparing the entire functions *t → f* (*V*^*a*=1^) with *t → f* (*V*^*a*=0^) ^18^, rather than ascribing treatment effects based on the hazard ratio at points in time. ^19,20^ The basic parameter in a multi-state model is the transition-specific intensity or hazard ^21^ *α*_*ij*_ between all pairs of states *i* and *j*,

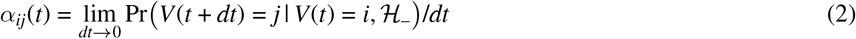

where *ℋ*_–_ is a filtration on (0, *t*], denoting the history of the process. If the only conditioning is on the current state (*ℋ*_–_ does not affect the intensity), then the transition has the Markov property. A multi-state model is said to be a Markov model when all transition intensities are Markov. The marginal hazard function describes the instantaneous risk of reaching a state at time *t*, given that it has not yet been reached, and can be given as

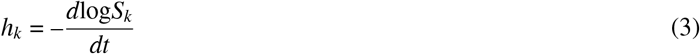

where *S*_*k*_ is the survival function for the time to reach state *k*. When competing events may prevent the outcome from occurring, the time to the event of interest may be formally infinite for some individuals, and the causal parameter requires careful interpretation. In this work, the functional of interest is the marginal hazard of time to first entry of a specific state (the state represents the occurrence of a clinical outcome of interest), when all patients begin in the same starting state. We interpret this parameter as a cause-specific hazard for the outcome. We give the expression for the marginal survival function for a multi-state model, which can be converted to the marginal hazard using equation 3, as

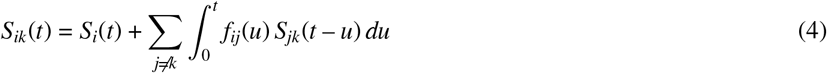

where *S*_*ik*_ is the marginal survival function starting in state *i*, i.e. not reaching state *k* by time *t*. This function is not a simple function of hazards (unless all transition hazards obey the Markov assumption). The first term in equation 4 relates to staying in the initial state up to *t*, and the second term is a path-dependent sum over all states accessible from *i* (excluding *k*), with a convolution over *t* jumping to *j* at time *u* and surviving in *j* for the remaining time *t* – *u*. When transitions are reversible, then the *S*_*jk*_ terms are themselves a sequence of recursive convolutions. This analytical complexity makes it difficult to derived closed form expressions for the estimands and thus we use microsimulation to estimate the true value of the estimands.

The fundamental problem of causal inference requires three assumptions to hold in order for an observational study to be conceptualised as a conditionally randomised experiment in which associational parameters can be interpreted causally. The three assumptions are positivity, conditional exchangeability and consistency. We furthermore require independent censoring to hold in order for hazards (the fundamental quantity of interest) to be identifiable. We give formal definitions of these below.

### 3.1 Notation and assumptions

The primary time scale of interest is time since initial state entry, corresponding to simulation start, which we denote *t*_0_. Let *V*(*t*) denote the state occupied at time *t*. We let *A*(*t*) be a binary time-dependent indicator function denoting actual treatment status at time *t* (*A* = 1 is treated). *Y*^*a*^ is the distribution of the potential outcome assuming *A* = *a*, and *L* is a *p*–dimensional vector of time-dependent covariates. *a* and *l* represent specific values of *A* and *L*. Let *R*(*t*) represent the at-risk process such that

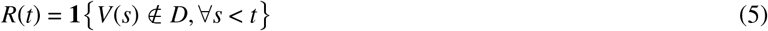

where *D* is the set of absorbing states. In words, the patient is at-risk of transitions if the patient has not reached an absorbing state prior to time *t*. We denote the full history of a variable with an overbar. In general, the history of treatment *Ā* (*t*) and covariates 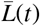 until time *t* may affect the potential outcomes. For a treatment with two possible values, a treatment strategy *g* in continuous time will be a sequence with possible values *a* ∈ (0, 1) at each time, for example, the always treat strategy is defined by *g* = (1, 1, …1). The data is generated from a multi-state model which operationalises a structural causal model. The structural model *ℳ* can be stated generally as

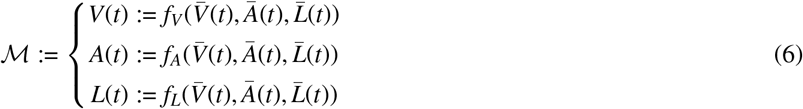

and the specific transition intensities are the structural equations in this model. We now describe the assumptions required for causal inference in our setting.

1. **Positivity:** Pr[*A* = *a*|*L* = *l*] > 0 (For all treatment strategies under consideration, the treatment rate should be non-zero.)
2. **Sequential conditional exchangeability:** *Y*^*a*^ ***⊥*** *A*|*L*
3. **Consistency:** *Y* = *Y*^*g*^ (the realised outcome is consistent with the potential outcome for the particular value of treatment received.)
4. **Modularity:** do 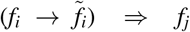 unchanged for all *j ≠i*. (an intervention on any one mechanism replaces only its corresponding structural equation, leaving all others invariant.)
5. **Independent censoring** Censoring at *C* is independent if

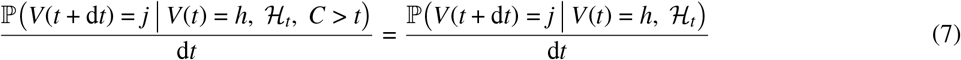

where *ℋ*_*t*_ is the history of the process up to *t*. The modularity assumption is required for interventions on outcome regression models to be valid, and is satisfied if the outcome model is correctly specified. Both consistency and modularity are generally satisfied in simulation studies when the estimand is obtained using micro-simulation, which is the case in this work.

## 4 SIMULATION AIM

The aim of the simulation study was to evaluate the performance of different estimation approaches under various datageneration scenarios relevant to deprescribing where a drug is abruptly stopped (i.e. not tapered). Independent and identically distributed life-history data were simulated from a structural causal model based on a continuous-time multi-state model. The parameters of the DGM were mainly based on hypothetical values, the most common choice, as per our recent review of the pharmacoepidemiology literature. ^22^. Our choice here was influenced by the highly conditional nature of the state-space in reversible non-Markov multi-state models and the lack of transition intensity estimates in the literature. If the process is reversible and non-Markov, the states visited in the sample path are not determined by the knowledge of the states occupied at visit times, unless extra information is available. ^23^ We modelled all-cause mortality as a state which was accessible from any other state, and therefore it was possible to use a real-world estimate for this aspect of the DGM. We pre-specified and pre-registered a protocol for this study which states the DGM parameters and methods. ^24^ The code is on Zenodo. ^25^

## 5 DATA GENERATING MECHANISM

We generated life histories independently for *n*_obs_ patients using a custom multi-state model with 5 states. The state space *v* represented the following health states

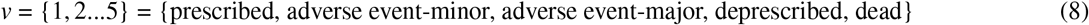

All patients begin in *V*(0) = 1. Conceptually, we define this as a period in which the patient has been prescribed (for a period of at least two years) and is fully adherent to their medication regimen (i.e., taking it regularly). We also assume full adherence throughout this study. The exposure value of each patient (prescribed/deprescribed) is part of the stochastic process. The transition structure of the multi-state model is shown in Figure 1. The transition-specific intensities from state *i* to *j* for person *k* are defined using multiplicative hazards of the form

**FIGURE 1.**
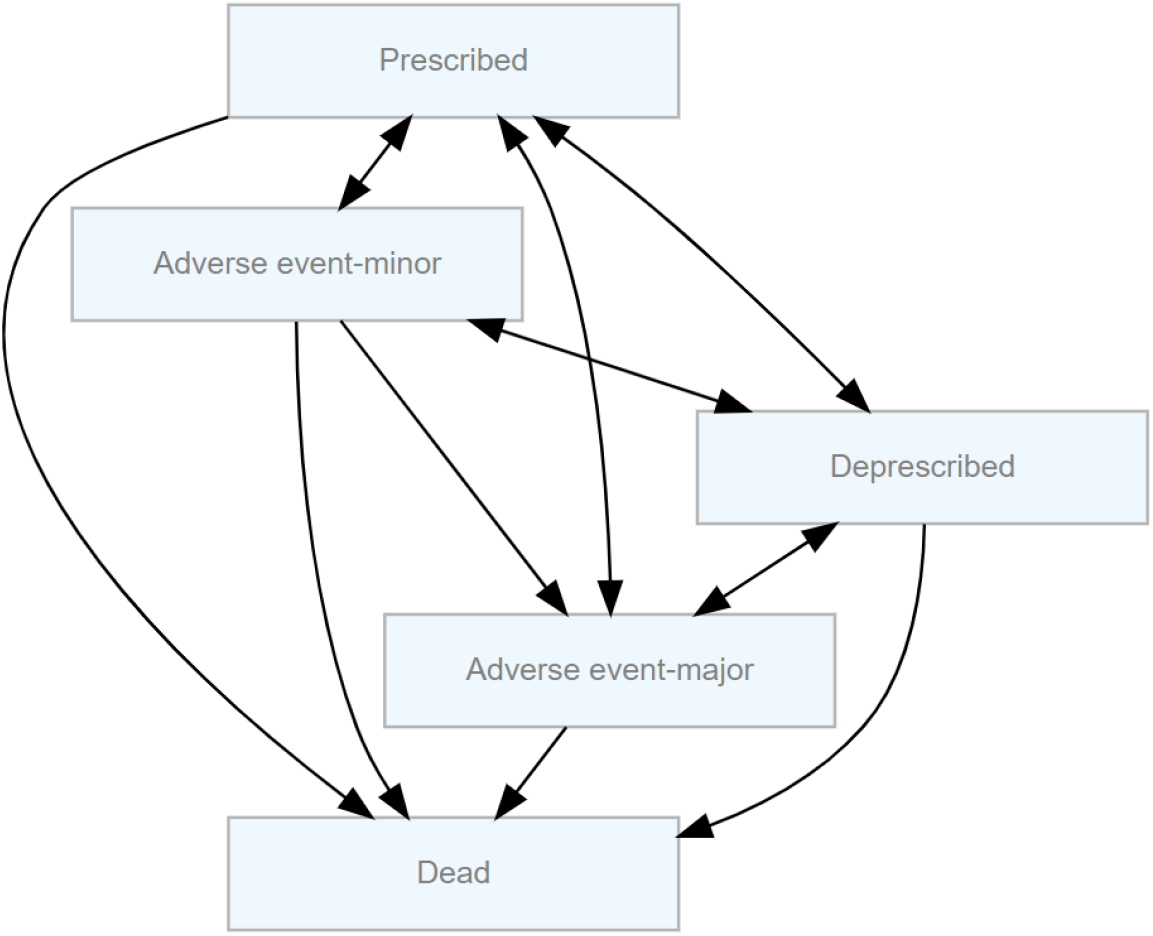
State transition structure of the multi-state model.

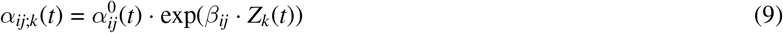

where 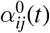 is the baseline hazard, *β*_*ij*_ is a 1 *× p* vector of coefficients, and *Z*_*k*_(*t*) is a *p ×* 1 vector of covariates. When the transition intensities depend on the past only through the current state and possibly time-fixed covariates, the multi-state process is a Markov process. Baseline hazards were defined using parametric functions, as shown in Table 2. The Weibull hazard rate was parametrised as

**TABLE 2.** Parameter values of baseline hazard rates.

| **Transition** | **Distribution** | **a (months<sup>−1</sup>)** | **b (months<sup>−1</sup>)** | **Shape (-)** | **Scale (months<sup>−1</sup>)** | **Source** |
|---|---|---|---|---|---|---|
| Prescribed to AE-major | Weibull | - | - | 1.00 | 5.00 × 10<sup>−3</sup> | CJ<sup>*a*</sup> |
| Prescribed to AE-minor | Weibull | - | - | 1.00 | 1.00 × 10<sup>−2</sup> | CJ |
| Prescribed to dead | Gompertz | 3.28 × 10<sup>−5</sup> | 7.53 × 10<sup>−3</sup> | - | - | LT<sup>*b*</sup> |
| Prescribed to deprescribed | Weibull | - | - | 1.00 | 2.96 × 10<sup>−3</sup> | RWE<sup>*c*</sup> |
| Deprescribed to prescribed | Weibull | - | - | 1.00 | 1.00 × 10<sup>−3</sup> | CJ |
| Deprescribed to dead | Gompertz | 3.28 × 10<sup>−5</sup> | 7.53 × 10<sup>−3</sup> | - | - | LT |
| Deprescribed to AE-minor | Weibull | - | - | 1.02 | 8.30 × 10<sup>−3</sup> | CJ |
| Deprescribed to AE-major | Weibull | - | - | 9.50 × 10<sup>−1</sup> | 8.30 × 10<sup>−3</sup> | CJ |
| AE-minor to prescribed | Weibull | - | - | 1.00 | 5.00 × 10<sup>−4</sup> | CJ |
| AE-minor to AE-major | Weibull | - | - | 1.00 | 1.00 × 10<sup>−2</sup> | CJ |
| AE-minor to deprescribed | Weibull | - | - | 1.00 | 5.92 × 10<sup>−3</sup> | CJ |
| AE-minor to dead | Gompertz | 3.28 × 10<sup>−5</sup> | 7.53 × 10<sup>−3</sup> | - | - | LT |
| AE-major to prescribed | Weibull | - | - | 1.00 | 1.00 × 10<sup>−4</sup> | CJ |
| AE-major to deprescribed | Weibull | - | - | 1.00 | 2.96 × 10<sup>−2</sup> | CJ |
| AE-major to dead | Gompertz | 3.28 × 10<sup>−5</sup> | 7.53 × 10<sup>−3</sup> | - | - | LT |

<sup>*a*</sup> CJ: Clinical judgement
<sup>*b*</sup> LT: EUROSTAT lifetables <sup>26</sup>
<sup>*c*</sup> RWE: Real world evidence <sup>27</sup>

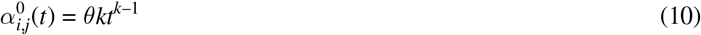

where *θ, k ∈* ℝ+ are the scale and shape parameters of the Weibull distribution. The Gompertz hazard rate was parametrised as

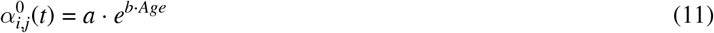

Where *a, b* and *Age* are the Gompertz scale and shape parameters and the patient’s age, respectively. We obtained realistic values of *a* and *b* from a parametric fit of the EU 27 average all-cause death hazard for 2022, truncated to ages 3 to 80. ^26^ (see supporting information Figure S1). This resulted in a mean life expectancy of approximately 81 years. We specify patient-specific covariates that induce heterogeneity in the intensities. Covariates are obtained from an initial multivariate distribution

*G* and their time-evolution was given by deterministic functions of *t* and past process history. All covariates are *V*-adapted processes, such that at any change of state, their function can be evaluated. Simulation of transitions used Monte-Carlo integration with nested root-finding. ^28^ (see simulation algorithm 1) The covariates are given in Table 3. The time-evolution of the covariates is shown in Table S1.

**TABLE 3.** Covariates and their roles, data types, time-dependency, sources, and examples.

| **Covariate** | **Role** | **Data type** | **Time-dependent** | **Source** | **Example** |
|---|---|---|---|---|---|
| *L*<sub>1</sub> | Baseline confounder | Continuous | Yes | External | Age |
| *L*<sub>2</sub> | Time-varying exposure | Binary | Yes | Internal | Deprescribed indicator |
| *L*<sub>3</sub> | Time-varying mediator | Ordinal | Yes | Internal | *N*<sub>adverse-events</sub> |
| *L*<sub>4</sub> | Time-varying confounder | Continuous | Yes | External | Comorbidity |
| *L*<sub>5</sub> | Effect modifier | Binary | No | External | Sex |
| *L*<sub>6</sub> | Indicator of deprescribing | Binary | Yes | Internal | Ever deprescribed indicator |

The parameters of *G* are given in Table S2 and the correlation matrix is given in equation 2 of the supporting information. The values of the coefficients *β*_*i,j*_ are shown in Table S3. All baseline transition intensities except for 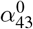 and 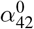 are defined on the simulation time scale *t* (clock-forward). We modify the two hazards above to be defined relative to the current start time *t*_*s*_ of the exposure process. Thus, the time scale used is *t* – *t*_*s*_. This is due to the exposure process being embedded as a state within the model.

### 5.1 Factor variables

The factor variables we systematically vary in the simulation are the population sample size *n*_obs_ and the 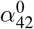 transition hazard *θ*_43_ (deprescribing to AE-major). The latter factor will be obtained using

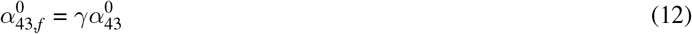

where 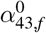 is the factor *f* hazard. All DGM factors are shown in Table 4.

**TABLE 4.** DGM factor levels.

| Factor | *γ* | HR (*t* = 1) | *n*<sub>obs</sub> |
|---|---|---|---|
| Factor 1 | 1 | 1.5770 | 2000 |
| Factor 2 | 0.9 | 1.4193 | 2000 |
| Factor 3 | 0.8 | 1.2616 | 2000 |
| Factor 4 | 0.5 | 0.7885 | 2000 |
| Factor 5 | 1 | 1.5770 | 1600 |
| Factor 6 | 0.9 | 1.4193 | 1600 |
| Factor 7 | 0.8 | 1.2616 | 1600 |
| Factor 8 | 0.5 | 0.7885 | 1600 |
| Factor 9 | 1 | 1.5770 | 1000 |
| Factor 10 | 0.9 | 1.4193 | 1000 |
| Factor 11 | 0.8 | 1.2616 | 1000 |
| Factor 12 | 0.5 | 0.7885 | 1000 |

### 5.2 Observable data from the DGM

Our data available for inference was intended to represent routinely collected healthcare data, since this study focusses on using the TTE framework to infer causal effects from observational data. We distinguish between two distinct types of observation that captures the underlying state of individuals in an observational setting; event- or symptom-driven visits and scheduled visits. ^29^ Event-driven visits occur due to some change in health state (e.g. worsening symptoms) and is influenced by the patients’ desire or need to see their provider. Scheduled visits are specified by the healthcare provider at a prior visit, usually for monitoring the starting/stopping of a treatment. Scheduled visits may thus be uneventful and not record any change in health state unless there was an incidental finding, whereas event-driven visits always record a change in health state, and the patient’s covariates may be measured. This study models event-driven visits only, resulting in multi-state data of the true health process, where each life-history is subject to an independently generated right-censoring time *C*. Changes of state are thus representative of visits, at which, the state and all values of covariates are known. We define the analysis set as the information available for inference, representing the observed EHR data. Let *ℱ* = {*ℱ*_*t*_ : *t ≥* 0} denote the full filtration generated by the multi-state data-generating mechanism. For each subject *i, ℱ*_*i*_(*t*) describes the set of {*V, L*} for all times up to t. The analysis filtration *ℱ*_*a*_, representing the information available to the analyst is then

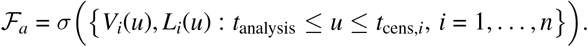

where *t*_analysis_ is some left truncation time and *t*_cens,*i*_ is an individual right censoring time. We chose *t*_analysis_ = 60 months and sample *t*_cens,*i*_ from an exponential distribution with rate parameter *λ* = 0.0002 months^-1^.

## 6 ESTIMANDS

In a review of target trial emulation studies in medicine, ^30^ the hazard ratio was the most commonly reported measure of effect. Thus, the hazard ratio was chosen as the target parameter in this study. Two clinically relevant estimands were defined with reference to the addendum on estimands and sensitivity analysis in clinical trials document ICH E9 R1. ^7^ The target parameter was the counterfactual hazard ratio corresponding to the time until first major adverse event (entry to state 3 in the model) for two distinct contrasts of treatment strategies. This parameter, which we term 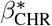 is a projection of the true (possibly time-varying) hazard ratio on to a proportional hazards (PH) working model ^31^, and is the solution to the following score equation

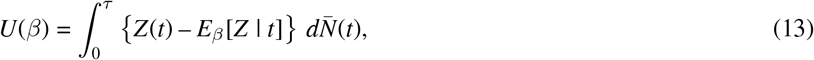

where *Z*(*t*) denotes the (possibly time-dependent) exposure process. The term 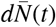 denotes the increment of the population event counting process, representing the infinitesimal probability mass of failure at time *t* under the counterfactual distribution. *E*_*β*_[*Z* | *t*] is the Cox model-implied risk set mean of the exposure process, defined as

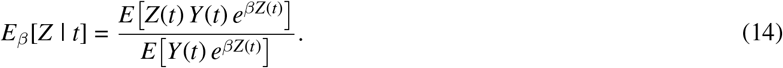

Here, *β* is the log-hazard ratio parameter indexing the proportional hazards working model

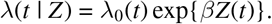

Finally, *U*(*β*) is the Cox partial likelihood score functional which defines the estimating equation for the target parameter via *E*[*U*(*β*)] = 0.

We now give the specific estimands used in the simulation.

### 6.1 Estimand 1

The first estimand compares *g*_1_ and *g*_2_, which are the treatments defined by specifying the initial state as 4 (deprescribed) versus 1 (prescribed) at *t*_analysis_ respectively. We term this the intention-to-treat estimand (ATE_*ITT*_ ).

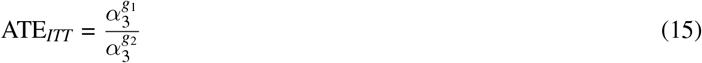

Figure 2 (a) describes the single-world intervention graph (SWIG). ^32,33^ for estimand 1.

**FIGURE 2.**
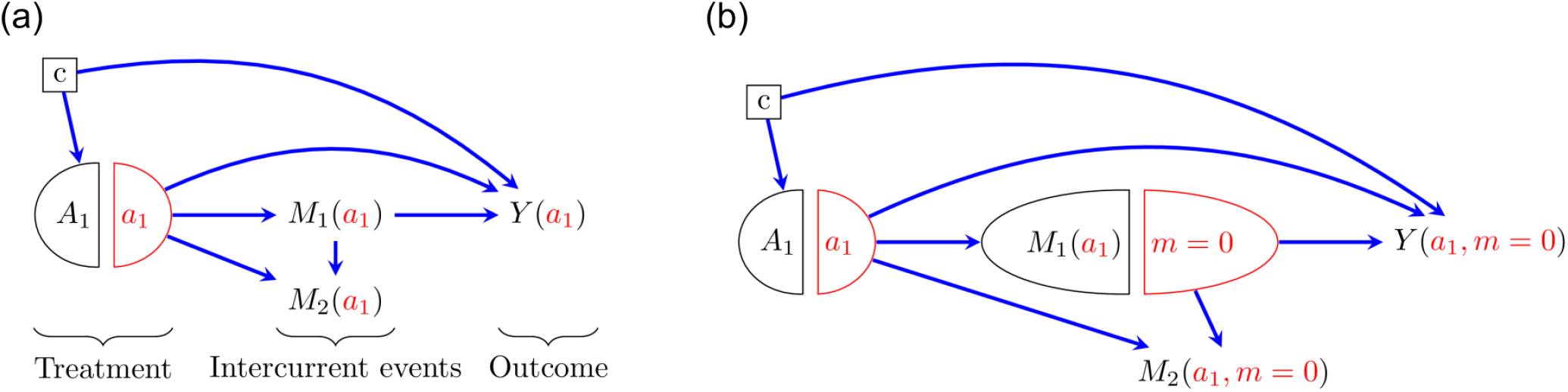
(a) SWIG for estimand 1. *M*_1_ is the occurrence of treatment switching after exposure (restarting of the medication of interest after deprescribing has occurred), *M*_2_ is the occurrence of death and *c* is the set of confounders. (b) SWIG for estimand 2.

### 6.2 Estimand 2

The second estimand compares *g*_3_ and *g*_4_, which correspond to always being in state 4 and always being in state 1 respectively. This was termed the per-protocol estimand (ATE_*PP*_).

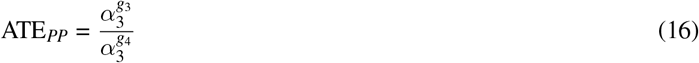

For the ITT estimand, the hazard rates are given as

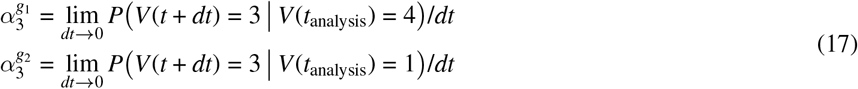

where we marginalise over the covariate distribution and condition only on state occupation at *t*_analysis_. For the PP estimand, the rates additionally condition on the exposure process remaining constant (at the value set at *t* = *t*_analysis_) using artificial censoring when treatment becomes inconsistent with this strategy. We also marginalise over the covariate distribution. Thus, the hazard rates are

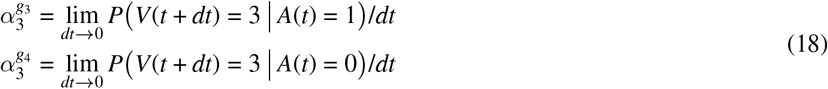

Figure 2 (b) shows the SWIG for estimand 2.

### 6.3 Defining the true estimand

The true estimand was defined as the causal hazard ratio of deprescribing versus continuing to be prescribed with the failure time being the time to the first major adverse event (*V* = 3). We both ignored and accounted for treatment deviation by defining an intention-to-treat (ITT) effect and per-protocol (PP) effect respectively. The true ITT effect was estimated from an oracle dataset which was generated using a hypothetical intervention characterised by setting the initial state occupied by the whole population at simulation time zero. This intervention relates to immediately deprescribing the whole population at time zero. Thus, two counterfactual datasets were generated, each having identical baseline covariates, however the exposed group had *A* = 1 and the unexposed group had *A* = 0. We termed the latter dataset the “natural course”, since it represents the natural sequence of events for our population, i.e. the population at *t*_0_ was defined to be those prescribed and without any episodes of prior deprescribing, whom may stochastically enter the deprescribed state over time. The dataset with A = 1 was termed the intervened dataset. All inference was performed on the natural course dataset only. The estimand was computed using Monte-Carlo integration. ^34^

We note that in order to ensure positivity for the treatment strategies, the population at time zero of analysis should consist of people with both types of exposure status. Thus, all of the analyses begin at time *t*_start_ > *t*_0_. The choice of *t*_start_ affects the characteristics of the simulated population, for example by reducing the proportion of those patients alive and event free over time or by changing the proportion of exposed patients. To align the analysis and oracle populations in this study, the oracle dataset was modified to consist only of those patients who were alive and event-free within the natural course at *t*_start_, and their covariates were updated to those that would be measured at *t*_start_. This was possible since all covariates were adapted processes based on known deterministic functions.

The ITT estimand was determined using the natural course and intervened datasets. The PP estimand was obtained using two additional datasets with initial state intervention and modified transition intensities. A Cox proportional hazard model was fitted with the only covariate being the dataset indicator (exposed/unexposed) and the hazard ratio was obtained. We note that under a general non-proportional hazards DGM, the estimate obtained from the Cox partial likelihood estimator is not a time-averaged hazard ratio over the follow up period, the so-called average hazard ratio (aHR) is a separate quantity ^35^. We note that the parameter obtained from the Cox partial likelihood estimate depends on the censoring distribution. ^36,31^ Thus, the estimand in this simulation study can be viewed as a projection of the true (time-varying) HR onto a PH working model. A natural solution is to include more general models, however as the PH assumption is used in both oracle and inference estimation, it will not affect the performance measures. We thus assume that the true estimand is the value obtained from the partial likelihood estimator.

Furthermore, in this study, we assumed prescribing data and relevant outcome data (e.g., when someone is prescribed a medicine and the diagnosis of an adverse event) is all that is available for inference. Dispensing data may also be available but we assume this is not the case in this study. In this case, an intention-to-treat effect can be defined based on prescribing status at a single point in time. Estimation of the ITT effect in the oracle dataset used the initial state occupation as the sole exposure status, regardless of the post-baseline status. Estimation of the PP effect used intervention on cause-specific hazards to set the hazard of treatment switching to zero. This consisted of setting all exposure specific transition hazards to zero, keeping all other hazards at the value defined by the natural course. This hypothetical intervention assumes that prevention of treatment switching is independent of potential outcomes (modularity assumption).

In summary the ITT estimand focused on a point-in-time exposure *Ā* = *A*(0) = *a* and the PP estimand focused on a sustained exposure over time *Ā*= *A*(*t*) = *a*. These strategies are deterministic static treatment strategies. For identifying the PP estimand, we make the assumption that treatment switching is independent of the hazard and artificially censor when this occurs. Thus, conditional on the relevant covariates, the counterfactual hazard for those who change exposure status is assumed equal to that of those who remain on their assigned treatment up to time *t*.

## 7 SIMULATION METHODS

We focused on the most common continuous-time parametric and semi-parametric likelihood-based methods. Non-parametric methods based on hazard estimation require some transformation (e.g., smoothing) to summarise the risk measure ^21^ and so these methods were not assessed. We used two common approaches to correct for the lack of exchangeability, inverse probability of treatment weighting and standardisation using parametric outcome regression. Matching was not considered as it may exclude some eligible individuals from the analysis, depending on the matching approach used. The methods implemented were naive Cox regression, inverse probability of treatment weighted (IPTW) Cox regression, the naive sequential Cox method, IPTW weighted sequential Cox regression, extended Cox regression and a parametric multi-state model based on the g-formula.

### 7.1 Crude Cox proportional hazards model

The Cox model is the most ubiquitous modelling method used for failure times in epidemiology. The semi-parametric form of the Cox model provides useful flexibility in terms of the absolute risk of events, while often making the assumption of proportional hazards for modelling simplicity. It has been stated that the Cox model (and other models providing contrasts of counterfactual hazards) do not generally allow causal interpretation of the contrasts. ^37^. This is because the hazard function conditions on survival to time *t*, but the population at *t* may be affected by prior effects of treatment, or non-treatment effects (selection effects), and therefore is not identical to the randomised population. This is a feature of period-specific contrasts, which can be ignored when taking a functional view of hazards over time. ^18^ The functional view of hazards discourages point-in-time interpretations due to the above selection effects, but also prevents any interpretation of hazards as being direct effects.

The marginal hazard function for the Cox proportional hazards model is defined as

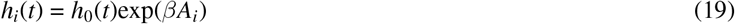

where *h*_0_(*t*) is an unspecified baseline hazard, *β* is the log hazard ratio between the exposed and unexposed groups, and *A*_*i*_ is the treatment indicator (being in the deprescribed state at *t*_analysis_). In this model the time zero is *t*_analysis_ and the event time is the first entry to state 3. As the estimand is a marginal treatment effect, there are no further covariates included in the regression model. Since the exposure groups are simulated using an intensity process which has confounding as a feature, this model is not expected to provide consistent estimates of the causal hazard ratio.

### 7.2 Marginal structural model - IPTW

The backdoor method can be used to ensure that treatment is independent of measured confounders *Y ⊥ T*|*L*, using the inverse probability of treatment weights (IPTW). Application of IPTW weights in a marginal Cox model is one implementation of estimating a marginal structural model (MSM). ^38^ A Cox MSM is a *direct* model for counterfactual hazards. The general form of a Cox MSM is

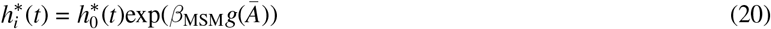

where 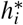 is the counterfactual hazard a patient would experience if their treatment history was set to 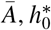 is the baseline hazard someone would would experience if their treatment history was set to *g*(*Ā*) = 0, *g* is a function of the history of treatment and covariates and exp(*β*_MSM_) is the ratio of these two counterfactual hazards. When fitting the MSM, *g* is pre-specified according to the treatment strategy. For both estimands in the simulation, *g* was the value of observed treatment status at baseline. For estimand 2, we artificially censor individuals when their treatment status deviates from baseline.

Estimation of the parameters of an MSM requires ensuring exchangeability, which is typically performed using weighting strategies by employing a parametric model for exposure. This involves weighting subject-specific observations by a value proportional to 1/*e*(*L*) in the Cox model, where *e*(*L*) = ℙ (*A* = *a*|*L* = *l*) is known as the propensity score. This method can provide consistent estimates of the causal parameters of equation 20 under exchangeability, positivity and consistency. Further inclusion of the marginal treatment probability in the numerator of the weights provides stabilisation of the estimation procedure, and typically results in greater efficiency. We use a logistic regression model to calculate the propensity score using all available confounders

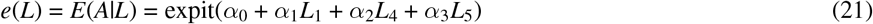

with the subject-specific weights computed as

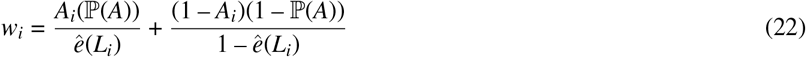

The *w*_*i*_ are used to weight each observation in a Cox model using the coxph function in R.

### 7.3 Crude sequential Cox model

The analysis of times-to-events may be applied in a sequential trial approach, whereby a number of cohort entry times are chosen from which to measure follow-up. ^39^ Exposure is assessed prior to this time such that each time zero mimics the randomisation time of a hypothetical trial. This approach will generally lead to greater data utilisation compared to using a single calendar-based cohort entry time. since a patient can contribute to more than one trial if they meet the inclusion criteria more than once. This approach was implemented for both estimands using an approach similar to the method in section 7.1. The major difference is that in this method, several different baseline times were used to assess inclusion and treatment status at pre-specified intervals.

There are two time-related quantities affecting the interpretation of this method; the interval between entry times and the follow-up time of each trial. The trial interval Δ_trial_ is the difference in calendar time between time zero of one trial and the next. In a multi-state setting with event-driven visit times, a fixed interval will likely not align exactly with observation times, thus for each patient the time zero of the *i*^th^ trial was calculated as

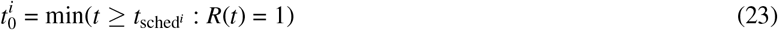

where the *i*^th^ scheduled trial time is given as

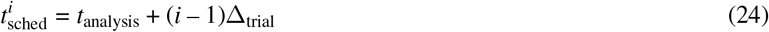

In the simulation Δ_trial_ was chosen as 20 months and the maximum scheduled trial start time as 100 months. In real world settings, the interval should be chosen based on the frequency of the events of interest and exposure changes and other information, such as the treatment dynamics. After defining the sequence of trial start times, the analysis proceeds identically to section 7.1 by constructing multiple risk sets and pooling all trial data to obtain the marginal hazard ratio.

One consideration in a closed-cohort setting such as that in this work is the effect of covariate drift on the construction of the risk sets. Necessarily some time-dependent covariates (e.g., age) will vary systematically, and if these have a non-zero effect on the outcome, then it may be preferable to stratify analysis by this variable or use trial-specific random effects. In the simulation, we expect covariate drift to have a negligible effect on the pooled HR estimate due to the magnitude of time-dependent effect modification.

### 7.4 IPTW sequential Cox model

The analysis of the sequential Cox model in section 7.3 was also performed using IPTW weighting in order to estimate the parameters of the MSM corresponding to equation 20.

### 7.5 Extended Cox model

A different variation of the proportional hazards model was also implemented, based on the extended Cox model. The hazard function for this model is given as

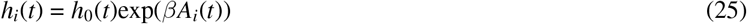

which is identical to equation 19 other than the relaxation of the exposure process to now be a possibly time-dependent variable. The expression above makes no restriction on when *A*(*t*) is evaluated, and so it is typically evaluated when exposure changes (the hazard is conditional on the “current” value of exposure). This model then implicitly assumes that the hazard obeys the Markov property. ^40^ Interpretation of *β* is now slightly different compared to equation 19. The estimate is now the hazard ratio of the time to the event of interest given current exposure status, also known as the partial transition rate or partly-conditional rate. ^41^ Since the exposure process *A*(*t*) is an endogenous covariate, the hazard is conditional on a random and therefore unpredictable process. ^42^ Thus, from a causal perspective, the estimate is difficult to interpret because the intervention is not well defined, i.e. consistency is not immediately ensured.

### 7.6 Parametric g-formula

An alternative approach to estimate causal effects is to parametrically model the conditional expectation of the outcome and standardise to the observed covariate distribution. The parametric g-formula is one such implementation which enables the estimation of counterfactual quantities. There are two algebraically equivalent but different versions of this method, the noniterative conditional expectation (NICE) and iterative conditional expectation (ICE) versions. ^43^ We focus on the NICE version in this study. For a general treatment strategy *a*, with baseline covariates *w*, the marginal functional of a multi-state process can be restated using the law of total expectation

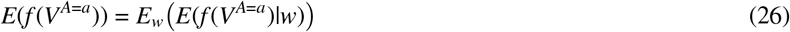

When the sequential exchangeability assumption holds

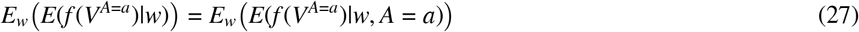

while consistency grants

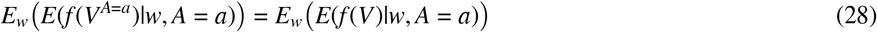

where the right-side of equation 28 is now a function of observed data only.

In order to estimate the ATE we can substitute model-based estimates of *E*(*f* (*V*)|*w, A* = *a*) into equation 1 and compute the empirical average over independent observations

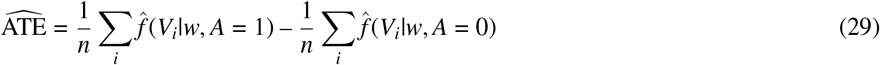

The above estimate is known as the g-computation formula, and when fully parametric models are used, it is termed the parametric g-formula. This method requires positivity and correct specification of the model for the conditional expectation for valid inference. Thus, under certain assumptions, the g-formula relates a process summary functional to the expectation of conditional functionals over the observed distribution of covariates.

In this study the choice of parametric model was a Markov multi-state model with identical transition structure to the true DGM and rates modulated by baseline covariates. Therefore, by definition, the parametric model was misspecified since the true DGM incorporated non-Markov transition intensities. This was a decision based on practicality in real data analysis scenarios, where the true parametric form of several conditional hazard models would not be known. Assuming Markovianity is also ubiquitous in multivariate survival settings. We fitted the Markov model on the unfiltered dataset (i.e. since simulation time zero), since strict positivity is not required when exposure is one of the states in a multi-state model. Since the only simulated censoring was random right censoring, each transition hazard could be estimated independently by treating competing event times (all state entry times) and administrative censoring times as right censoring. We estimated and simulated all hazards using the `flexsurv` package.

After obtaining estimates for the model, we simulated counterfactual data for each estimand as follows.

**Estimand 1: Intention-to-treat** This was a static non-sustained strategy.

1. Intervene on the initial state, setting *V*(0) to 1 and generate a dataset by simulating events until the absorbing state was reached. Generate a similar dataset by setting *V*(0) to 4.
2. These two datasets were then combined and used to calculate the summary functional of interest by estimating the marginal hazard ratio from a model with dataset indicator as the only covariate.

**Estimand 2: Per-protocol** This was a static sustained strategy using a joint intervention that combines:

1. The intervention on the initial state as in Estimand 1
2. Interventions on the cause-specific hazards affecting the embedded exposure process (For *V*(0)=1, the intervention sets: *α*_*n*4_(*t*) = 0 *∀n* and for *V*(0)=4, the intervention sets: *α*_*n*1_(*t*) = 0 *∀n*).
3. The target parameter was then obtained similarly to estimand 1.

Standard errors were obtained using the non-parametric bootstrap procedure with 20 samples. We acknowledge that this is a low number of repetitions. Obtaining variance estimates for the g-formula (and therefore its sequential extension) estimators is a known problem due to computational complexity. ^44^ This work was similarly limited in this regard, however qualitative convergence of standard errors was obtained with the number of bootstrap samples used.

### 7.7 Clone-censor weighting

The final method implemented was the increasingly popular clone-censor weight (CCW) method. ^45,4^ CCW was originally introduced in order to estimate the parameters of a marginal structural model of which the target was a dynamic treatment regime. ^46^ This was motivated by the fact that a person’s treatment status may temporarily be consistent with multiple treatment strategies. Thus, replication of a sample (cloning) ensures that the modelling accounts for the multiple types of exposure received. Another application of CCW is when the treatment is a random static regime, such as “treat within 10 days of baseline”. In the context of failure-time outcomes, artificial censoring may be used to handle intercurrent events. CCW consists of the application of cloning individuals at baseline, artificially censoring when the treatment status is no longer consistent with that of the clone and weighting the sample by the inverse probability of being censored to ensure that artificial censoring is conditionally independent of the outcome. In this work, CCW was implemented using data augmentation and standard Cox regression.

The data augmentation consisted of choosing a time window for assessing the start of treatment status. In this study we chose 90 months. Each patient was then cloned at baseline in to a group representing each treatment status (prescribed/deprescribed). If, by the end of the window, a person had entered in to the deprescribed state, the patient in the prescribed group would have their follow-up artificially censored at that time. If a patient did not enter the deprescribed state by the end of the interval, those in that group would have their follow-up censored at the end of the interval.

## 8 PERFORMANCE MEASURES

All performance measures were computed using the rsimsum package. ^47^ The bias was estimated from all included simulation replicates according to:

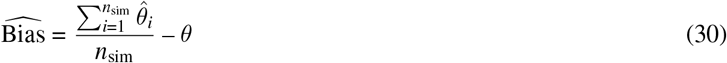

where *θ* is the true treatment effect and 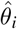 is the effect estimate from replication *i*. The Monte-Carlo standard error (MCSE) of the bias was estimated with

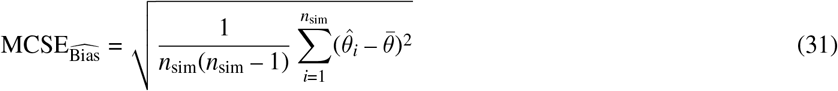

where 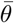 is the mean of 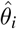. The coverage of the 95% confidence intervals was estimated using

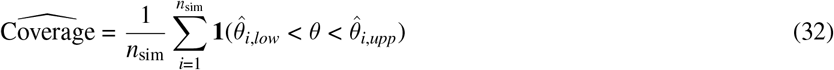

where **1**{*·*} is the indicator function, *θ*_*i,low*_ and *θ*_*i,upp*_ are the lower and upper boundary of the i^th^ intervals. The MCSE of coverage was estimated using

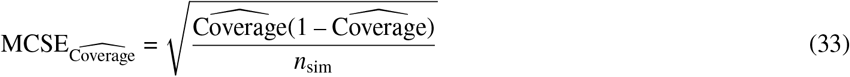

In this work, performance measures are reported as 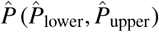, where 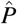 denotes the point estimate of the performance measure, and 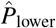 and 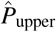 denote the lower and upper bounds of the corresponding 95% confidence interval of the measure, respectively.

## 9 RESULTS

In Figure S2 an alluvial plot of the first five state transitions in the natural course dataset (*n*_sim_ = 1) for the base case factor are shown. The number of failed estimation attempts is shown in Figure S3. The corresponding missing estimates only occurred for the TD-Cox method, and there were at most 3 (0.75%) missing estimates per factor. We assumed these estimates were ignorable in the analysis of the performance measures. Thus, a complete case analysis per method and per factor was used.

In Figure 3 a heatmap of the point estimate of the bias is shown for the all DGM factors. The DGM scenarios all had timedependent confounding and non-zero true effect sizes. The *γ* factor impacts the true value of the estimand because it varies the Weibull scale parameter of the hazard of interest. The *γ* value of 0.5 had an induced HR closest to the null at *t* = 1. Although *γ* scales the baseline value of the *α*_43_ transition hazard, since the target estimand is based on non-proportional hazards, this does not scale the estimand linearly. The new value depends on the full functional form of the hazards. The other factor controls the number of observations used for estimation, effectively simulating how varying sample sizes affect the performance measures. It can be seen that, for the ITT estimand, the marginal structural model and IPTW sequential trials methods were the least biased and the clone-censor-weight method, crude sequential trials, crude Cox, parametric g-formula and time-dependent Cox methods all had large amounts of bias. For the ITT estimand, as *γ* decreased, the bias estimates for the CCW and TD-Cox methods increased in magnitude. There was no trend with respect to the null effect. It can be seen that for the PP estimand, the clone-censor-weight method, crude sequential trials, crude Cox, parametric g-formula were all highly biased, with the CCW and g-formula methods having bias that varied significantly with respect to *γ*. We give tabulated values including confidence intervals for all performance measures in Tables S4-S7 of the supporting information. For the two least biased methods and across both estimands, there appeared to be no trend in the value of bias versus the scale parameter coefficient *γ*. Figure 4 shows the interval estimate of the coverage for each method, factor and estimand. It can be seen that the marginal structural model and IPTW sequential trials methods were the only methods with calibrated confidence intervals that achieved nominal coverage. All other methods had anti-conservative coverage. In Figure S4, the performance measures for the sensitivity analysis are shown, where the number of observations was 10,000.

**FIGURE 3.**
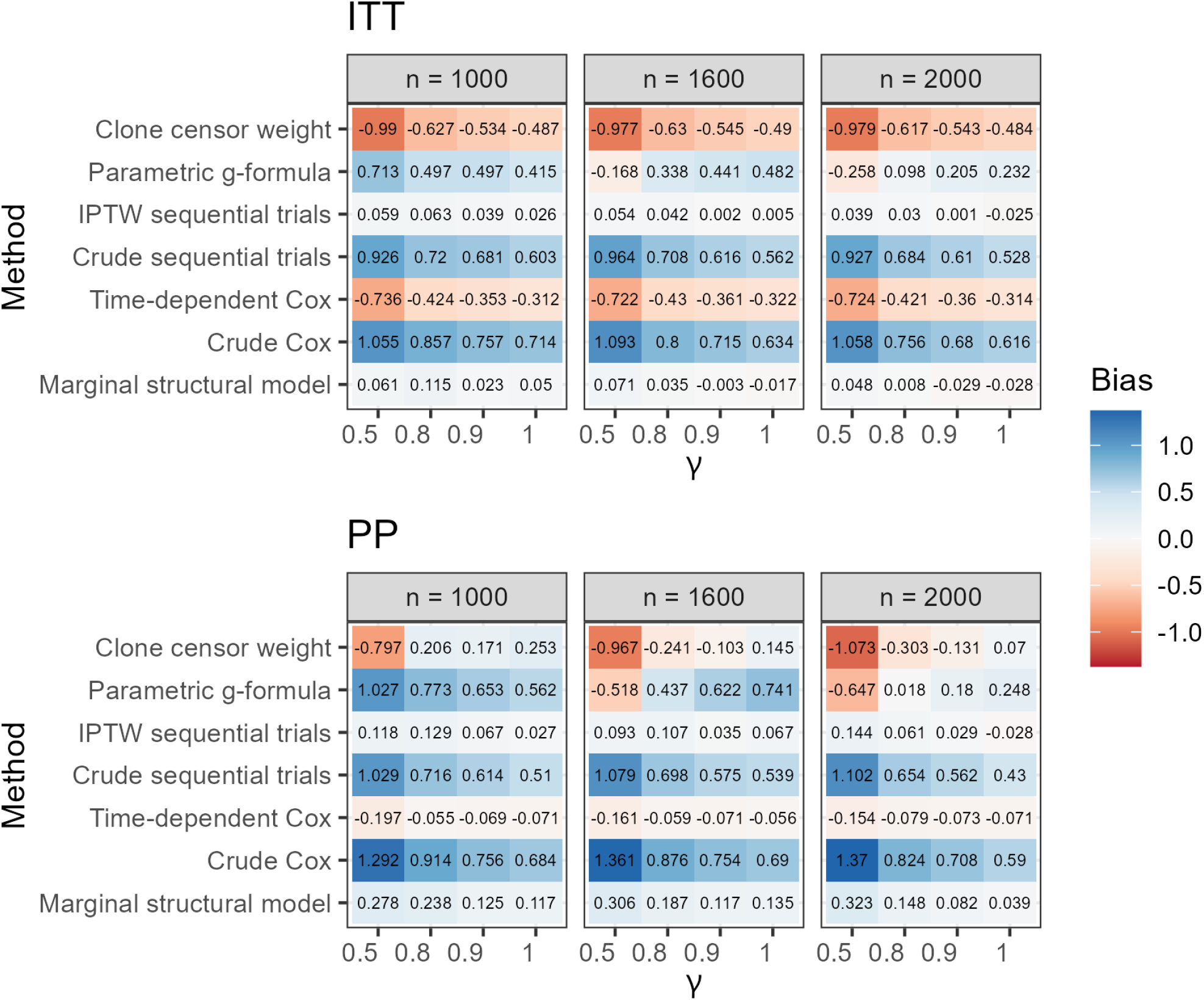
Point estimate of the bias for each method and DGM factor. ITT: intention-to-treat estimand, PP: per-protocol estimand. *γ* is a multiplier of the state 4 to state 3 baseline hazard (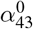). *n* corresponds to the value of *n*_obs_. The base case was *n*_obs_ = 2000 and *γ* = 1.

**FIGURE 4.**
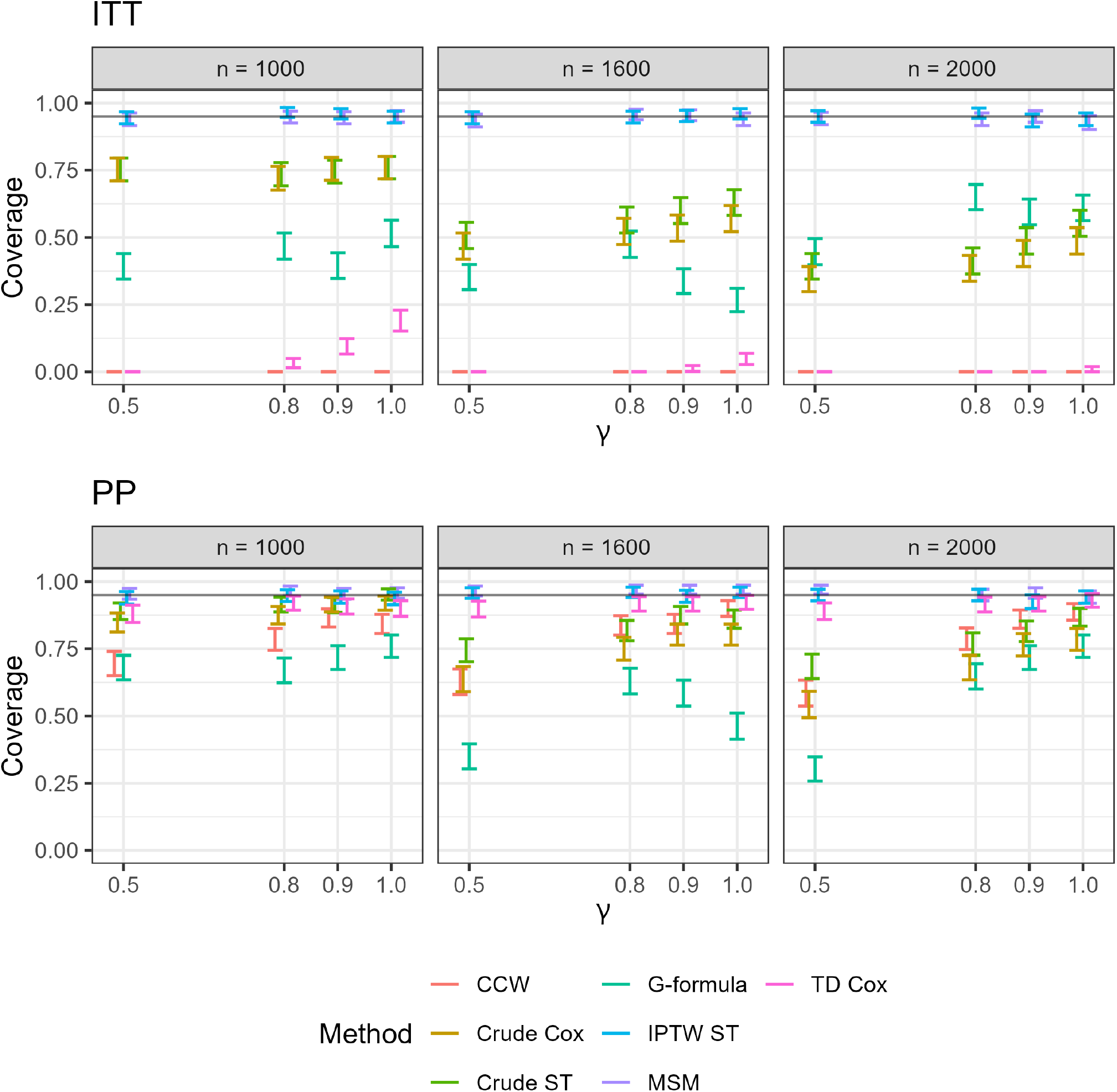
Interval estimate (95 % confidence interval) of the probability that confidence intevals at the 5 % level contain the true value of the estimand for each method and DGM factor. ITT: intention-to-treat estimand, PP: per-protocol estimand. Horizontal line indicates the nominal coverage level. *n* corresponds to the value of *n*_obs_.

## 10 DISCUSSION

This simulation study has found meaningful differences in bias and coverage across different methods to estimate the causal effect of medication deprescribing using the target trial emulation framework. Each of the performance measures will be discussed in detail in separate sections below.

### 10.1 Bias

While bias is inherently quantitative, it is useful to categorise bias according to either the known or likely source. In simulation studies, the source of bias can be differentiated based on knowledge of the DGM and estimators used. An analogue of bias is the concept of statistical error in single samples. Kawahara *et al*. ^48^ distinguished between three sources of statistical error (the difference between a sample estimate and its true value); (1) estimand error, (2) structural error and (3) error due to random noise. This categorisation can be used to similarly describe bias sources. However, since bias is the long-run difference between the estimate and its true value, random noise by definition should not contribute to bias. In a simulation study with a finite number of simulation repetitions, however, the performance measures are subject to Monte Carlo error, making it difficult to distinguish residual random variation from systematic bias. We assume that the number of simulation repetitions is sufficiently large that the Monte Carlo standard errors of the performance measures are negligible relative to the differences in bias being investigated. For all DGM factors, estimands and methods, there were no outliers that had a disproportionate effect on the bias.

We first focus on estimand error, also known as estimand misalignment. This source of bias is likely to occur when the research question is either ill-defined and/or the translation into a statistical estimand is not correct. In target trial emulation, clearly specifying the question and the estimand is paramount to the methodology and heavily encouraged. ^49,50^ In our example target trial, the question concerns the effect of stopping a drug immediately versus continuing, which describes a single-point exposure. The question also suggests that the summary measure is the hazard ratio. Without further information, this might suggest that proportional hazards are assumed since this is the most common approach to obtaining a unidimensional hazard-based parameter. If we did not intend on making such assumption, then the stated estimand in Table 1 would be misaligned with the research question. Choice of the estimator may also contribute to error when non-parametric identifying assumptions are not fulfilled (structural bias), either through lack of specification or inappropriate estimators.

For example, in this study, the CCW method is misaligned with the estimand of interest. This is due to the nature of the estimator, which (in its most common implementation) targets the causal effect of treatment strategies with random, finite-horizon initiation periods. Examples of such strategies being “begin treatment within 30 days of time zero”. This is in contrast to the treatment strategies defined in the target trial, which are defined deterministically at time zero. This is the major source of bias for the ITT estimand, since, as per the treatment policy strategy there is no deviation from the strategy and no artificial censoring (for treatment deviation) is necessary. The estimate collapses to unity regardless of the data because there is no variation in the outcome between groups. This also results in empirical standard errors of zero since there is no variation in the estimate with respect to *n*_sim_. For the PP estimand, while there is artificial censoring upon treatment deviation, there is no obvious choice for the length of the initiation period (since it is effectively zero for the estimand). Choosing zero would result in all estimates collapsing to unity. For informative purposes, we chose this arbitrarily as 90 months. Bias can be expected in this case because there are underlying effect modifiers that vary over time, e.g. age, that may attenuate or increase the risk of the outcome at later times, which was the case in this work. Hence, the CCW method was heavily biased for the ITT estimand, with a value of -0.484 (-0.484, -0.484) but was unbiased for the base case and the PP estimand with a value of 0.070 (-0.029, 0.170). However, the bias was heavily dependent on the number of observations *n*_obs_ and the lack of bias for the *n*_obs_ = 2000 factor was coincidental.

The other method which suffers from estimand error is the time-dependent Cox (TD-Cox) model. As mentioned in section 7.5, this model targets the partial transition rate. This estimator estimates a parameter from a common multi-state model, the three-state reversible model with 1 absorbing state, also known as the healthy-illness-death model with recovery. The estimand obtained in the TD-Cox model corresponds to the ratio of the partial transition rates of illness to death and healthy to death. The standard Cox model incorporating the proportional hazards assumption either implies a constant hazard ratio over time or a projection of the true hazard ratio on to a constant in a Cox partial likelihood sense, which is dependent on the censoring mechanism and length of follow-up. When using a TD-Cox model where the hazard may depend on the current value of exposure, then the estimate obtained further depends on the exposure process, which may change at different times for different people. When the underlying multi-state process is Markov, the ratio of partial transition rates equals the ratio of transition intensities. In this simulation, since the hazards were non-Markov, the estimator resulting from the TD-Cox model does not target the correct estimand. The observed bias is thus a result of estimand error for both the ITT estimand, with a value of - 0.314 (-0.320, -0.308), and PP estimand, with a bias of -0.071 (-0.092, -0.050). The TD-Cox model incorporated adjustment for confounding using time-updated inverse-probability of treatment weights and so structural bias was not an issue for this method. The estimates obtained were identical for both estimands, since it uses the current value of exposure. Therefore differences in the bias between estimands is only due to differences in the true value of the estimand. It can be seen that the bias for the PP estimand is relatively small. This is expected because the ratio of partial transition rates closely resembles the true hazard ratio, for this estimand. For per-protocol estimands, only information from exposure consistent with that at baseline contributes to the estimate. The small amount of bias is due to time-dependent effect modification that is present.

We now discuss the presence of structural bias, due to the failure of identifying conditions required for inference. The identifying conditions always required are the three basic causal inference assumptions of positivity, exchangeability and consistency. When estimands include time-to-event outcomes, independent censoring is also required. Since administrative censoring in this study was generated independently for all patients, this assumption is fulfilled. As mentioned in section 3.1, consistency is fulfilled in this study by design. Modularity is only required for models employing outcome regression, such as the g-formula. Positivity and exchangeability are thus the two potential causes of structural bias for the methods. It should be noted that the clone-censor-weight method is particularly invulnerable to structural bias, since exchangeability at the index time is guaranteed and the random nature of the treatment strategy provides variation in the allowable initiation time. However, as already mentioned, the random nature of the exposure is also potentially undesirable when answering certain research questions that have a deterministic exposure, such as “begin treatment at time zero”.

For both estimands, the two methods which do not account for confounding, the crude Cox and crude sequential trials methods are severely biased. Confounding was due to both baseline values of covariates and the time-updated values of covariates affecting the risk of the outcome and exposure. For the ITT estimand, the bias in the crude Cox model is entirely due to nonexchangeability, since positivity was ensured by the estimand definition. The crude sequential trials method enjoys greater precision due to the use of a larger number of samples, however since it does not account for confounding it was biased as a result.

In the base case DGM, the parametric g-formula had a bias of 0.232 (0.213, 0.250) for the ITT estimand and 0.248 (0.215, 0.280) for the PP estimand. This was expected due to the parametric form of the outcome model used. Since the deprescribed to AE-major event hazard was generated from a Weibull distribution, this resulted in violation of the Markov assumption. Thus, the cause-specific regression model was misspecified for this transition. The result was severely biased away from the null for all other DGM factors except factors 8 and 4. In deprescribing, time-varying hazards of the outcome of interest are expected. Reasons for this are that the physiological effects of stopping a medication may not be immediate and there may be short-term withdrawal effects soon after deprescribing that cease after some time. Certain benefits of deprescribing, such as increased focus and sleep quality ^51^ may also take some time to occur.

The interval estimate of bias for the marginal structural model and IPTW sequential trials methods included zero for four DGM factors and for both estimands. However the point estimate was close to zero for all factors. There was no apparent trend in the bias with respect to the number of observations or the true effect size. For the large value of *n*_obs_, the IPTW sequential trials method had a bias that included zero, while the marginal structural model had a nonzero bias for both estimands respectively. However, the upper estimate for the ITT estimand was -0.003 (less than 1 % relative bias), and we expect the estimate would tend to zero as *n*_sim_ increases.

Interestingly, the PP estimate for the methods that accounted for non-exchangeability was also slightly biased due to practical violations of positivity, as seen in Figure 3. The IPTW sequential trials method had a bias of -0.028 (-0.062, 0.005) for the base case. After inspecting individual replicate cumulative incidence curves, the cause of this bias was attributed as follows. In order to estimate the causal effect in the hypothetical scenario where subjects do not switch treatment status, each method artificially censors subjects at the point of a treatment deviation intercurrent event. Consequently, within some simulation replicates, few or no individuals remained at risk of the outcome at longer follow-up times. When no individuals remain at risk, the outcome process cannot be estimated beyond that time point. When only a small number of patients remained at risk, estimation also became unstable. This can be particularly problematic for estimands based on the hazard ratio, which depends on the composition of the risk sets over time. When the true hazard ratio varies over time, this truncated follow-up will induce a different estimate to the one where no censoring occurs due to the aforementioned dependence on the follow-up time. Marginal risk estimands such as the risk difference are also affected by insufficient information at later follow-up times, but do not depend on the evolving risk set composition as does a hazard ratio. When using sequential trials approaches, there is thus a need to ensure positivity in all trials, to prevent estimation failure and ensure correct inference. Within empirical studies, this may be achieved by using information on propensity scores to alter the dataset using truncation. However, modifying the data in this way has the potential unwanted effect of changing the population of interest.

### 10.2 Coverage

The coverage of each method reflects the joint performance of a method’s point estimation and variance estimation procedures. Confidence interval coverage is expected to be close to the nominal level when the estimator is approximately unbiased and the estimated standard errors accurately reflect the true sampling variability. The methods with large bias all had anti-conservative coverage, as expected. Even though all methods, where possible, used robust standard errors, the bias due to estimand and structural sources led to nominal coverage not being obtained. The CCW method had zero coverage for the ITT estimand for all DGM factors since the standard errors were zero for all replicates. This was due to the estimand definition as explained in section 10.1. For the PP estimand, censoring at treatment deviation from baseline led to a coverage probability of 0.888 (0.857, 0.918). The TD-Cox method had extremely low coverage of 0.010 (0.000, 0.020) for the base case and ITT estimand. The coverage for the PP estimand was 0.930 (0.905, 0.955). While this is unexpected due to the estimand misalignment, the coverage decreased accordingly with the increase in bias, as *γ* decreased. As such, the nominal coverage for this particular factor was coincidental. This highlights the importance of examining several performance measures in a simulation study, as well as several factors in the DGM.

A time-dependent Cox model is particularly tempting for scenarios where patients who experience improvement in symptoms are more likely to discontinue treatment, because treatment status changes frequently. This method may utilise more information compared to a Cox model using baseline exposure status only, and thus precision in the estimate may be higher. However, these results show that such a model may target a fundamentally different estimand from the effect of a well-specified deprescribing strategy. Consequently, a TTE using TD-Cox model could answer a different question from the one clinicians are interested in, while still producing apparently precise estimates due to the small standard errors it provides.

The coverage for the crude Cox model for the ITT estimand was 0.488 (0.439, 0.536) and for the PP estimand was 0.785 (0.745, 0.825). Coverage for the both estimands had a trend that was positively correlated with the *γ* parameter, and negatively correlated with *n*_obs_. The crude sequential trials method had a coverage of 0.552 (0.504, 0.601) for the ITT estimand and 0.868 (0.834, 0.901) for the PP estimand. The bias present in both of these methods was the source of low coverage, since the relative error in the model standard error was negligible.

The parametric g-formula had coverage of 0.610 (0.562, 0.658) for the ITT estimand and 0.760 (0.718, 0.802) for the PP estimand. Both bias and inaccuracy in the standard errors contributed to the observed anti-conservative coverage. The model-based standard errors differed from the empirical standard errors by approximately 30%, likely reflecting the relatively small number of bootstrap samples used to estimate uncertainty. We acknowledge this as a limitation of the study, as inaccurate standard error estimates may have affected confidence interval coverage. However, substantial bias was also present, as shown in Figure 3, indicating that the poor coverage was not solely attributable to variance estimation. Consequently, our conclusion that the implementation of a multi-state Markov model for time-varying hazards produced substantial bias remains unchanged. The poor performance observed for the parametric g-formula should not be interpreted as a limitation of the g-formula itself, but rather of the outcome model used to implement it. In this study, transition probabilities were estimated using a multi-state Markov model, which assumes that future transitions depend only on the current state and covariate history through the specified hazard model. When the underlying data-generating mechanism exhibits complex time-varying hazards, misspecification of the transition process may lead to biased estimates of intervention effects. Future research could investigate more flexible approaches for modelling transition hazards within the g-formula framework, including semi-Markov models, flexible parametric survival models, spline-based hazard models, or machine learning approaches. Such methods may better accommodate non-Markovian behaviour and complex hazard trajectories while retaining the ability of the g-formula to estimate causal effects under dynamic treatment strategies.

The marginal structural model had a coverage of 0.927 (0.902,0.953) for the ITT estimand and 0.943 (0.920, 0.965) for the PP estimand. The nominal coverage was achieved as expected due to all identifying assumptions being met. The IPTW sequential trials method had a coverage of 0.940 (0.917, 0.963) for the ITT estimand and 0.943 (0.920, 0.965) for the PP estimand. Although practical positivity violations introduced a small amount of bias for the PP estimand, this bias was insufficient to meaningfully affect confidence interval coverage, which remained close to the nominal level. There was no observed trend in the point estimate of coverage with respect to any DGM factors. This is expected for the type of factors varied in the study, since they had little impact on structural bias, and given that the methods were correctly specified and their identifying assumptions satisfied.

The results demonstrate that the methods which appropriately accounted for the confounding structure and targeted the estimand of interest provided approximately nominal confidence interval coverage for the effects of deprescribing interventions in the presence of time-varying confounding. In contrast, methods affected by structural biases, estimand misalignment, or outcome-model misspecification in the case of the parametric g-formula produced confidence intervals that failed to contain the true effect of the intervention. These findings suggest that when analysing observational deprescribing data, obtaining valid confidence intervals depend as much on selecting an appropriate causal framework and estimand as on accurate estimation of standard errors and correct modelling choices.

## 11 CONCLUSIONS

This study evaluated the performance of several methods commonly used in target trial emulation for estimating the effects of deprescribing interventions from observational data, in the context of deprescribing a medication as the intervention of interest. By generating data from a multi-state model with time-varying confounding, recurrent events, treatment switching, and non-Markov outcome processes, the study provided a realistic setting in which both methodological and practical challenges encountered in deprescribing research could be assessed.

The definition of a primary estimand is crucial for correct interpretation of clinical trials and observational studies emulating clinical trials. In this study, it was shown that within the target trial emulation framework, estimator choice can have a drastic impact on the performance of methods in terms of the ability to estimate the true value of an estimand as well as quantify the uncertainty in the estimate. While explicit specification of estimators is difficult without some knowledge of the data structure, it can be a vital tool in preventing failed estimation (through positivity violations) or misleading inference (through bias). Among the methods evaluated, the marginal structural model and inverse-probability-weighted sequential trials approach consistently demonstrated the best overall performance. Both methods produced negligible bias and achieved close-to-nominal confidence interval coverage across the range of data-generating mechanism factors investigated. Small departures from unbiasedness for the per-protocol estimand were observed due to practical positivity violations arising from artificial censoring at treatment deviations, but these effects were modest and had little impact on statistical inference. The use of multiple index times within the sequential trials method can improve precision since a larger sample size is obtained compared to using a single index time. The choice of IPTW ST versus an MSM should be based on the potential for covariate drift. This occurs when time-dependent effect modifiers systematically vary with respect to the time scale on which the trials are defined. For example, within closed cohort settings, the distribution of age and disease severity may be systematically different between early and later trials when using calendar time as the time scale. Alternative designs defining sequential trials based on age bands rather than calendar time may mitigate this. Such covariate drift shifts the population in which the causal effect is defined. If this is not consistent with the research question, then using a single index date (e.g., by employing an MSM) or estimating causal effects stratified by the index time may be required. These findings suggest that methods based on inverse-probability weighting provide a robust framework for estimating both intention-to-treat and per-protocol effects of deprescribing interventions when the required identifying assumptions are plausible.

The study also highlighted the importance of outcome-model specification when using outcome-regression approaches. Although the g-formula is theoretically capable of estimating the target estimand under the same identifying assumptions (since it is algebraically equivalent to the inverse weighting method), the implementation based on a multi-state Markov model performed poorly when the underlying hazards were non-Markov and varied over time. This suggests that future applications of the g-formula to deprescribing research may benefit from more flexible approaches for modelling transition hazards and event histories, when such behaviour is expected. The primary advantage of the parametric g-formula compared to weighting-based estimators is the ability to extrapolate over sparse regions of the covariate space, which can solve issues relating to positivity violations. This is however only possible when the conditional regressions are correctly specified.

Several limitations should be acknowledged. The simulation scenarios considered a specific multi-state structure and a limited set of confounding mechanisms, treatment processes, and outcome dynamics. Performance may differ under alternative data-generating mechanisms, particularly in settings with stronger positivity violations, measurement error, unmeasured confounding, or more complex treatment strategies involving longitudinal and/or dynamic regimens. Future work may examine the performance of the methods tested herein under these scenarios. We also did not systematically evaluate computational complexity of the methods, which is an important part of empirical studies. In addition, uncertainty for the g-formula was estimated using a limited number of bootstrap samples, which may have contributed to inaccuracies in variance estimation. Consequently, the findings should be interpreted as evidence for the scenarios studied rather than as absolute rankings of performance.

The pre-specification of several different estimands using various strategies to account for intercurrent events can provide clinicians with a hierarchy of evidence on the effects of varying levels of interventions. For example, the effect of initiating deprescribing using a treatment policy estimand may be useful for indicating whether or not it is effective to initiate deprescribing for a certain population. Other estimands based on hypothetical or while-on-treatment intercurrent event strategies may be used to infer the average treatment effect of deprescribing in a population over the whole course of the treatment (e.g., without restarting the medicine or using other rescue therapies), which may be more relevant for patients.

Overall, this work demonstrates that target trial emulation can be a valuable framework for estimating the effects of deprescribing interventions using observational data, but only when careful attention is paid to estimand specification, confounding adjustment, and model selection. In particular, methods that target the correct causal effect of interest and appropriately account for time-varying confounding were able to recover the true effects with minimal bias and approximately nominal confidence interval coverage. These findings provide practical guidance for researchers designing observational studies of deprescribing and highlight the importance of considering estimand alignment alongside traditional concerns regarding confounding and model misspecification. Currently, there are limited recommendations relating to deprescribing interventions ^52^ partly owing to limited high-certainty evidence on the benefits and harms of deprescribing. ^53^ The application of methods that account for the above complexities within observational research studies on the effects of deprescribing will support generation of less biased and more robust evidence on benefits and harms which may support future trial conduct and inform policy and practice.

## Data Availability

All code is available on Zenodo.

https://doi.org/10.5281/zenodo.22663632

## ACKNOWLEDGMENTS

None

