## Supplementary information for "Evaluating methods to quantify the effect of deprescribing a medication on patient clinical outcomes using a simulation study"

### SUPPORTING INFORMATION

### S1.1 | Gompertz fit from empirical life tables

The resulting parametric fit of the EU 27 average death hazard for ages 3 to 80 is shown below.

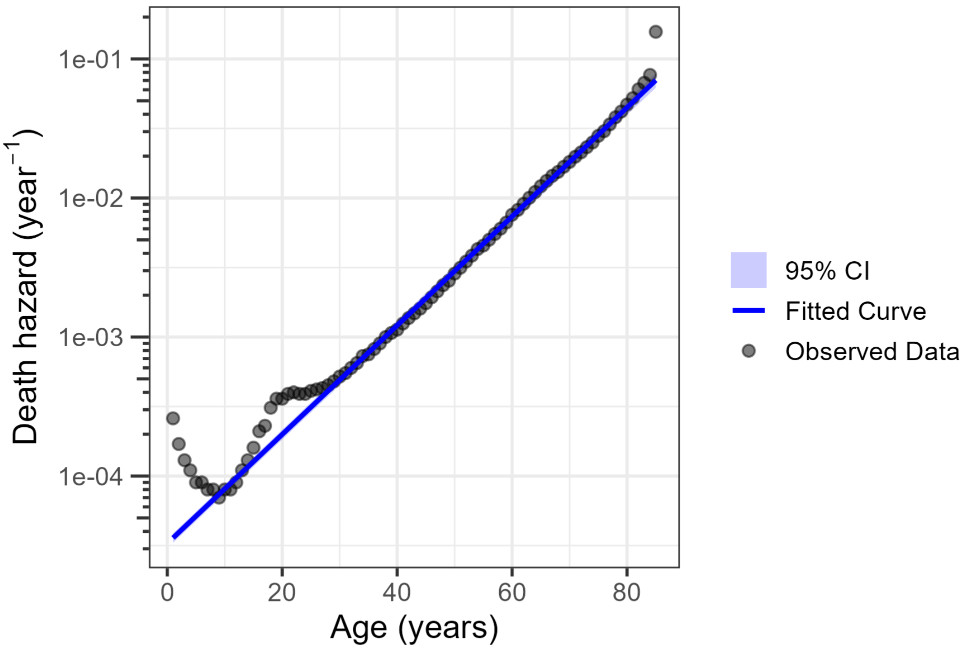


**FIGURE S1** Gompertz death hazard fitting results.

### S1.2 | DGM parameters

The algorithm for generating the data using a continuous multi-state process is given below.

**Algorithm 1** Continuous-Time DGM

**for** $i = 1, \ldots, n_{\text{obs}}$ **do**
  $v_i \leftarrow v_{\text{init}}$, $t \leftarrow 0$ ▷ initial state and time
  $L_i(0) \sim \mathcal{G}$ ▷ baseline covariates
  **while** $v_i \neq 5$ **do**
    Draw $u \sim \text{Uniform}(0, 1)$
    $\tau \leftarrow F^{-1}_{\text{all-cause}}(u \mid \mathcal{F}_t)$ ▷ holding time via inverse CDF
    $t \leftarrow t + \tau$
    $v \sim \text{Multinomial}\big(1; P(v \mid \mathcal{F}_t)\big)$ ▷ next state
    $L_i(t) \leftarrow \mathbf{X}(\mathcal{F}_t, \mathcal{G}, t)$ ▷ updated covariates
  **end while**
  Draw $\tau_c \sim \exp(\lambda_c)$
  $\mathcal{F}_a \leftarrow \sigma\big(\{V_i(s), L_i(s) : t_{\text{analysis}} \leq s \leq \tau_c\}\big)$ ▷ stopped filtration at the censoring time
**end for**

$\mathbf{X} = (X_1, \ldots, X_6)$ is the vector of covariate update functions, defined in TableS1. Since individual patients may not be at risk at $t_{\text{analysis}}$, we restrict the analysis to those who would be at risk at this time using a second simulation dataset starting from $t_{\text{analysis}}$, setting their baseline covariates to the value they would have at that time, under the natural course.

**TABLE S1** Covariate dynamics in the DGM

| Covariate | Update Rule |
|---|---|
| $X_1(t)$ | $\begin{bmatrix}1 & 0 & 0\end{bmatrix} \mathcal{G} + t$ |
| $X_2(t)$ | $0 + \mathbf{1}\{V(t) = 4, L_2(t_-) = 0\} - \mathbf{1}\{V(t) = 1, L_2(t_-) = 1\}$ |
| $X_3(t)$ | $\sum_{0<u\leq t} \mathbf{1}\{V(u_-) \neq 3,\ V(u) = 3\}$ |
| $X_4(t)$ | $\begin{bmatrix}0 & 1 & 0\end{bmatrix} \mathcal{G}(1 + 0.001t)$ |
| $X_5(t)$ | $\begin{bmatrix}0 & 0 & 1\end{bmatrix} \mathcal{G}$ |
| $X_6(t)$ | $0 + \mathbf{1}\{V(t) = 4\}$ |

*Note*: $\mathbf{1}\{\cdot\}$ denotes the indicator function, i.e., it equals 1 if the condition inside is true and 0 otherwise.

The distribution $\mathcal{G}$ represents the non-zero baseline covariates which are obtained from $\mathcal{G} = \begin{bmatrix}G_1 & G_2 & G_3\end{bmatrix}^{\mathrm{T}}$. $\mathcal{G}$ is thus a 3-dimensional mixed (Gaussian-Bernoulli) distribution with

$$\begin{aligned} G_1 &\sim \mathbf{N}(\mu_1, \sigma_1^2) \\ G_2 &\sim \mathbf{N}(\mu_2, \sigma_2^2) \\ G_3 &\sim \Pr(G_3 = 1) = p \end{aligned} \tag{1}$$

The parameters of the mixed distribution is given in table S2.

**TABLE S2** Parameters for non-zero baseline covariates.

| **Parameter** | **Value** |
|---|---|
| $\mu_1$ | 50 |
| $\mu_2$ | 0 |
| $\sigma_1^2$ | 10 |
| $\sigma_2^2$ | 1 |
| $p$ | 0.5 |

The target correlation matrix was specified as

$$\mathrm{Corr}(\mathcal{G}) = \begin{bmatrix} 1 & 0.2 & 0.2 \\ 0.2 & 1 & 0.2 \\ 0.2 & 0.2 & 1 \end{bmatrix} \tag{2}$$

The effects of covariates on the transition intensities is shown below

**TABLE S3** Covariate effects by transition intensity

| Covariate name | Age $X_1$ | Comorbidity $X_2$ | Sex $X_3$ | $N_{events}$ $X_4$ | Ever deprescribed $X_5$ | Deprescribed $X_6$ |
|---|---|---|---|---|---|---|
| Prescribed to AE-major | 0.00 | 0.00 | 0.00 | 0.00 | 0.00 | 0.00 |
| Prescribed to AE-minor | 0.00 | 0.00 | 0.00 | 0.00 | 0.00 | 0.00 |
| Prescribed to dead | 0.00 | 0.00 | 0.00 | 0.00 | 0.00 | 0.00 |
| Prescribed to deprescribed | 0.01 | 1.00 | 0.00 | 0.01 | -1.00 | 0.00 |
| Deprescribed to prescribed | 0.00 | 0.00 | 0.00 | 0.00 | 0.00 | 0.00 |
| Deprescribed to dead | 0.00 | 0.00 | 0.00 | 0.00 | 0.00 | 0.00 |
| Deprescribed to AE-minor | 0.00 | 0.00 | 0.00 | 0.00 | 0.00 | 0.00 |
| Deprescribed to AE-major | 0.01 | 1.00 | 1.00 | 0.01 | 0.00 | 0.00 |
| AE-minor to prescribed | 0.00 | 0.00 | 0.00 | 0.00 | 0.00 | 0.00 |
| AE-minor to AE-major | 0.00 | 0.00 | 0.00 | 0.00 | 0.00 | 0.00 |
| AE-minor to deprescribed | 0.00 | 0.00 | 0.00 | 0.00 | 0.00 | 0.00 |
| AE-minor to dead | 0.00 | 0.00 | 0.00 | 0.00 | 0.00 | 0.00 |
| AE-major to prescribed | 0.00 | 0.00 | 0.00 | 0.00 | 0.00 | 0.00 |
| AE-major to deprescribed | 0.00 | 0.00 | 0.00 | 0.00 | 0.00 | 0.00 |
| AE-major to dead | 0.00 | 0.00 | 0.00 | 0.00 | 0.00 | 0.00 |

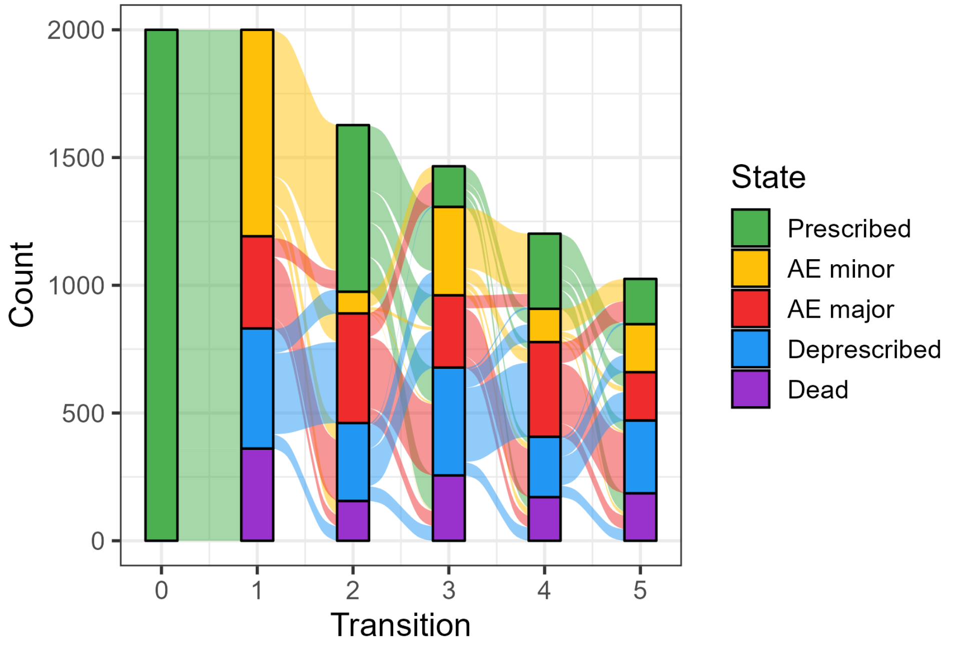


**FIGURE S2** Alluvial plot of the first five state transitions (all patients begin in the prescribed state).

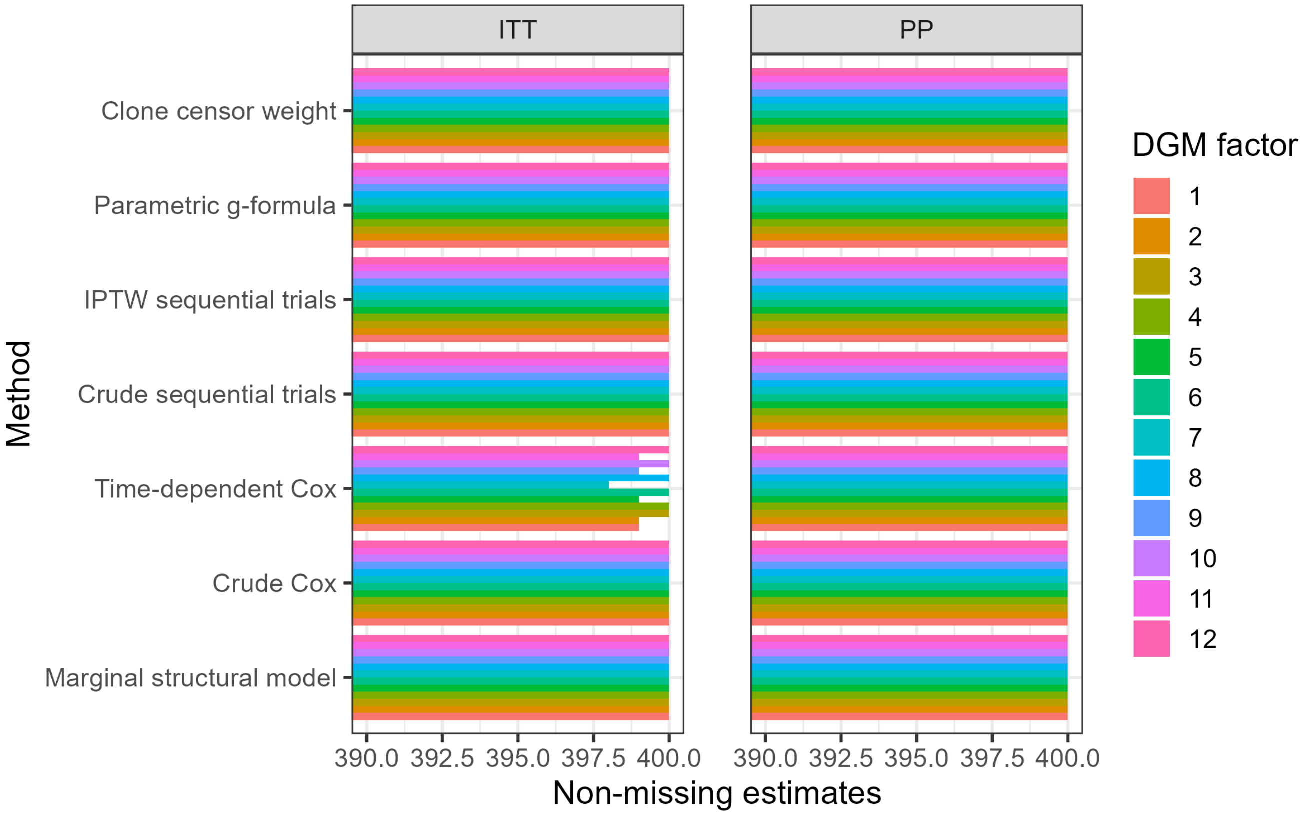


**FIGURE S3** Number of non-missing estimate values for all DGM factors and both estimands.

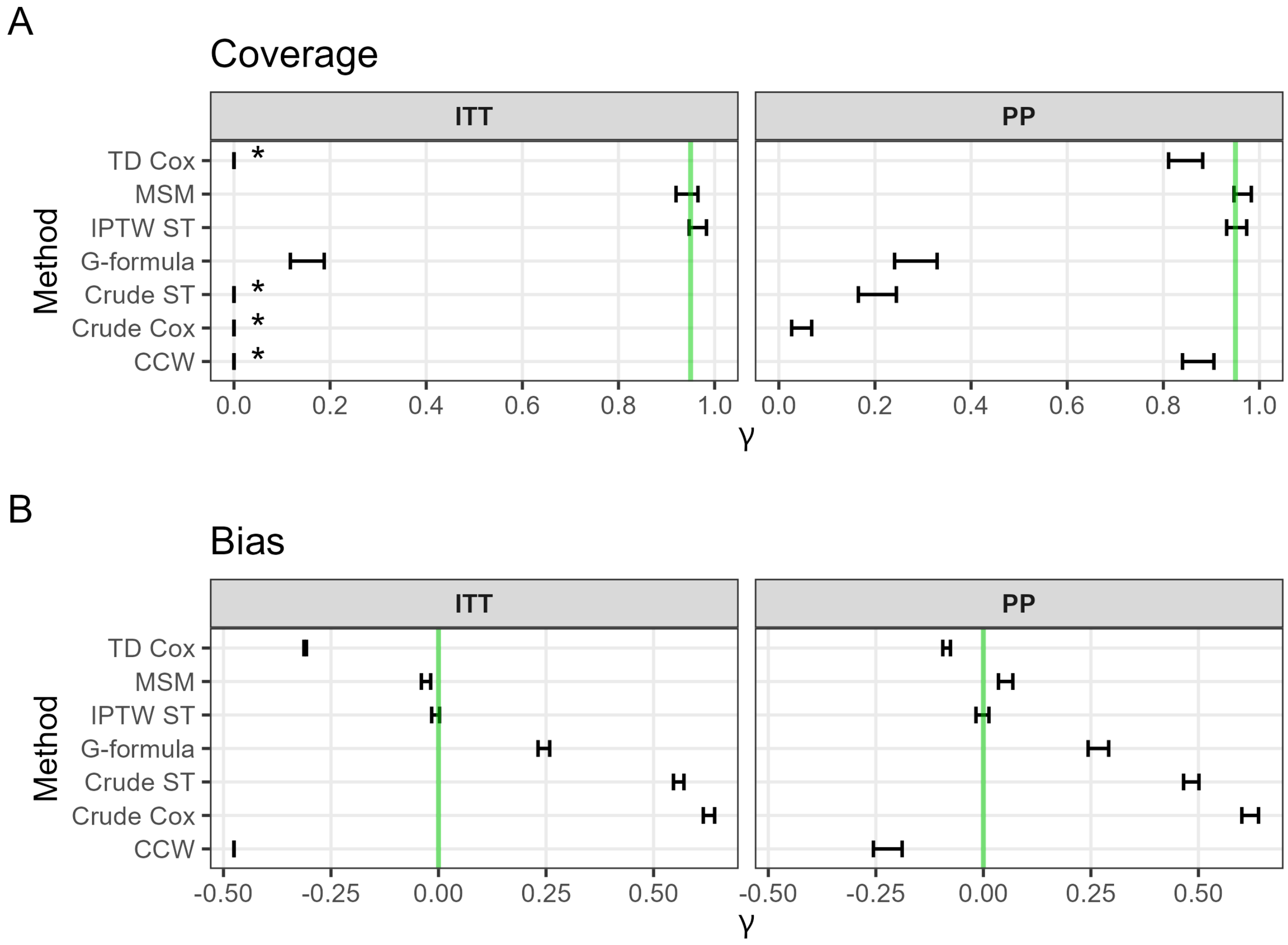


**FIGURE S4** 95% confidence interval estimate of the coverage ($\alpha$ = 5%) (a) and bias (b) for $n_{\text{obs}}$ = 10000 with the base DGM parameters. The green line corresponds to nominal coverage and zero bias respectively for each plot. (Asterisks indicate an upper value of zero for the interval).

**TABLE S4** Coverage: ITT estimand

| | CCW | | | G-formula | | | MSM | | | Crude ST | | | IPTW ST | | | TD Cox | | | Crude Cox | | |
|---|---|---|---|---|---|---|---|---|---|---|---|---|---|---|---|---|---|---|---|---|---|
| Factor | Est | Lower | Upper | Est | Lower | Upper | Est | Lower | Upper | Est | Lower | Upper | Est | Lower | Upper | Est | Lower | Upper | Est | Lower | Upper |
| 1 | 0 | 0 | 0 | 0.610 | 0.562 | 0.658 | 0.927 | 0.902 | 0.953 | 0.552 | 0.504 | 0.601 | 0.940 | 0.917 | 0.963 | 0.010 | 0.000 | 0.020 | 0.488 | 0.439 | 0.536 |
| 2 | 0 | 0 | 0 | 0.595 | 0.547 | 0.643 | 0.950 | 0.929 | 0.971 | 0.488 | 0.439 | 0.536 | 0.935 | 0.911 | 0.959 | 0.000 | 0.000 | 0.000 | 0.440 | 0.391 | 0.489 |
| 3 | 0 | 0 | 0 | 0.650 | 0.603 | 0.697 | 0.940 | 0.917 | 0.963 | 0.413 | 0.364 | 0.461 | 0.963 | 0.944 | 0.981 | 0.000 | 0.000 | 0.000 | 0.385 | 0.337 | 0.433 |
| 4 | 0 | 0 | 0 | 0.448 | 0.399 | 0.496 | 0.943 | 0.920 | 0.965 | 0.392 | 0.345 | 0.440 | 0.950 | 0.929 | 0.971 | 0.000 | 0.000 | 0.000 | 0.345 | 0.298 | 0.392 |
| 5 | 0 | 0 | 0 | 0.268 | 0.224 | 0.311 | 0.940 | 0.917 | 0.963 | 0.630 | 0.583 | 0.677 | 0.960 | 0.941 | 0.979 | 0.048 | 0.027 | 0.069 | 0.570 | 0.521 | 0.619 |
| 6 | 0 | 0 | 0 | 0.338 | 0.291 | 0.384 | 0.955 | 0.935 | 0.975 | 0.600 | 0.552 | 0.648 | 0.953 | 0.932 | 0.973 | 0.013 | 0.002 | 0.023 | 0.535 | 0.486 | 0.584 |
| 7 | 0 | 0 | 0 | 0.475 | 0.426 | 0.524 | 0.958 | 0.938 | 0.977 | 0.565 | 0.516 | 0.614 | 0.948 | 0.926 | 0.969 | 0.000 | 0.000 | 0.000 | 0.522 | 0.474 | 0.571 |
| 8 | 0 | 0 | 0 | 0.352 | 0.306 | 0.399 | 0.935 | 0.911 | 0.959 | 0.508 | 0.459 | 0.556 | 0.945 | 0.923 | 0.967 | 0.000 | 0.000 | 0.000 | 0.468 | 0.419 | 0.516 |
| 9 | 0 | 0 | 0 | 0.515 | 0.466 | 0.564 | 0.950 | 0.929 | 0.971 | 0.760 | 0.718 | 0.802 | 0.948 | 0.926 | 0.969 | 0.190 | 0.152 | 0.229 | 0.760 | 0.718 | 0.802 |
| 10 | 0 | 0 | 0 | 0.395 | 0.347 | 0.443 | 0.945 | 0.923 | 0.967 | 0.745 | 0.702 | 0.788 | 0.960 | 0.941 | 0.979 | 0.095 | 0.066 | 0.124 | 0.755 | 0.713 | 0.797 |
| 11 | 0 | 0 | 0 | 0.468 | 0.419 | 0.516 | 0.948 | 0.926 | 0.969 | 0.735 | 0.692 | 0.778 | 0.965 | 0.947 | 0.983 | 0.033 | 0.015 | 0.050 | 0.720 | 0.676 | 0.764 |
| 12 | 0 | 0 | 0 | 0.392 | 0.345 | 0.440 | 0.940 | 0.917 | 0.963 | 0.753 | 0.710 | 0.795 | 0.945 | 0.923 | 0.967 | 0.000 | 0.000 | 0.000 | 0.753 | 0.710 | 0.795 |

**TABLE S5** Coverage: PP estimand

| | CCW | | | G-formula | | | MSM | | | Crude ST | | | IPTW ST | | | TD Cox | | | Crude Cox | | |
|---|---|---|---|---|---|---|---|---|---|---|---|---|---|---|---|---|---|---|---|---|---|
| Factor | Est | Lower | Upper | Est | Lower | Upper | Est | Lower | Upper | Est | Lower | Upper | Est | Lower | Upper | Est | Lower | Upper | Est | Lower | Upper |
| 1 | 0.888 | 0.857 | 0.918 | 0.760 | 0.718 | 0.802 | 0.943 | 0.920 | 0.965 | 0.868 | 0.834 | 0.901 | 0.943 | 0.920 | 0.965 | 0.930 | 0.905 | 0.955 | 0.785 | 0.745 | 0.825 |
| 2 | 0.860 | 0.826 | 0.894 | 0.718 | 0.673 | 0.762 | 0.958 | 0.938 | 0.977 | 0.815 | 0.777 | 0.853 | 0.925 | 0.899 | 0.951 | 0.917 | 0.891 | 0.944 | 0.765 | 0.723 | 0.807 |
| 3 | 0.787 | 0.747 | 0.828 | 0.647 | 0.601 | 0.694 | 0.950 | 0.929 | 0.971 | 0.768 | 0.726 | 0.809 | 0.950 | 0.929 | 0.971 | 0.915 | 0.888 | 0.942 | 0.680 | 0.634 | 0.726 |
| 4 | 0.585 | 0.537 | 0.633 | 0.302 | 0.257 | 0.348 | 0.970 | 0.953 | 0.987 | 0.685 | 0.639 | 0.731 | 0.950 | 0.929 | 0.971 | 0.890 | 0.859 | 0.921 | 0.542 | 0.494 | 0.591 |
| 5 | 0.900 | 0.871 | 0.929 | 0.462 | 0.414 | 0.511 | 0.970 | 0.953 | 0.987 | 0.860 | 0.826 | 0.894 | 0.960 | 0.941 | 0.979 | 0.922 | 0.896 | 0.949 | 0.802 | 0.763 | 0.842 |
| 6 | 0.843 | 0.807 | 0.878 | 0.585 | 0.537 | 0.633 | 0.970 | 0.953 | 0.987 | 0.875 | 0.843 | 0.907 | 0.945 | 0.923 | 0.967 | 0.917 | 0.891 | 0.944 | 0.802 | 0.763 | 0.842 |
| 7 | 0.838 | 0.801 | 0.874 | 0.630 | 0.583 | 0.677 | 0.970 | 0.953 | 0.987 | 0.818 | 0.780 | 0.855 | 0.960 | 0.941 | 0.979 | 0.917 | 0.891 | 0.944 | 0.750 | 0.708 | 0.792 |
| 8 | 0.628 | 0.580 | 0.675 | 0.350 | 0.303 | 0.397 | 0.965 | 0.947 | 0.983 | 0.745 | 0.702 | 0.788 | 0.958 | 0.938 | 0.977 | 0.897 | 0.868 | 0.927 | 0.638 | 0.590 | 0.685 |
| 9 | 0.843 | 0.807 | 0.878 | 0.760 | 0.718 | 0.802 | 0.958 | 0.938 | 0.977 | 0.953 | 0.932 | 0.973 | 0.938 | 0.914 | 0.961 | 0.900 | 0.871 | 0.929 | 0.920 | 0.893 | 0.947 |
| 10 | 0.865 | 0.832 | 0.898 | 0.718 | 0.673 | 0.762 | 0.955 | 0.935 | 0.975 | 0.912 | 0.885 | 0.940 | 0.943 | 0.920 | 0.965 | 0.907 | 0.879 | 0.936 | 0.915 | 0.888 | 0.942 |
| 11 | 0.785 | 0.745 | 0.825 | 0.670 | 0.624 | 0.716 | 0.965 | 0.947 | 0.983 | 0.915 | 0.888 | 0.942 | 0.948 | 0.926 | 0.969 | 0.920 | 0.893 | 0.947 | 0.875 | 0.843 | 0.907 |
| 12 | 0.695 | 0.650 | 0.740 | 0.680 | 0.634 | 0.726 | 0.955 | 0.935 | 0.975 | 0.890 | 0.859 | 0.921 | 0.940 | 0.917 | 0.963 | 0.880 | 0.848 | 0.912 | 0.848 | 0.812 | 0.883 |

**TABLE S6** Bias: ITT estimand

| | CCW | | | G-formula | | | MSM | | | Crude ST | | | IPTW ST | | | TD Cox | | | Crude Cox | | |
|---|---|---|---|---|---|---|---|---|---|---|---|---|---|---|---|---|---|---|---|---|---|
| Factor | Est | Lower | Upper | Est | Lower | Upper | Est | Lower | Upper | Est | Lower | Upper | Est | Lower | Upper | Est | Lower | Upper | Est | Lower | Upper |
| 1 | -0.484 | -0.484 | -0.484 | 0.232 | 0.213 | 0.250 | -0.028 | -0.053 | -0.003 | 0.528 | 0.500 | 0.556 | -0.025 | -0.046 | -0.004 | -0.314 | -0.320 | -0.308 | 0.616 | 0.585 | 0.647 |
| 2 | -0.543 | -0.543 | -0.543 | 0.205 | 0.181 | 0.228 | -0.029 | -0.055 | -0.003 | 0.610 | 0.580 | 0.641 | 0.001 | -0.022 | 0.024 | -0.360 | -0.366 | -0.355 | 0.680 | 0.647 | 0.712 |
| 3 | -0.617 | -0.617 | -0.617 | 0.098 | 0.070 | 0.126 | 0.008 | -0.019 | 0.035 | 0.684 | 0.651 | 0.717 | 0.030 | 0.006 | 0.054 | -0.421 | -0.426 | -0.415 | 0.756 | 0.722 | 0.791 |
| 4 | -0.979 | -0.979 | -0.979 | -0.258 | -0.279 | -0.237 | 0.048 | 0.009 | 0.086 | 0.927 | 0.887 | 0.968 | 0.039 | 0.009 | 0.069 | -0.724 | -0.730 | -0.718 | 1.058 | 1.012 | 1.103 |
| 5 | -0.490 | -0.490 | -0.490 | 0.482 | 0.460 | 0.504 | -0.017 | -0.044 | 0.010 | 0.562 | 0.531 | 0.592 | 0.005 | -0.018 | 0.028 | -0.322 | -0.329 | -0.315 | 0.634 | 0.602 | 0.667 |
| 6 | -0.545 | -0.545 | -0.545 | 0.441 | 0.413 | 0.469 | -0.003 | -0.032 | 0.026 | 0.616 | 0.585 | 0.647 | 0.002 | -0.023 | 0.027 | -0.361 | -0.368 | -0.354 | 0.715 | 0.681 | 0.750 |
| 7 | -0.630 | -0.630 | -0.630 | 0.338 | 0.306 | 0.370 | 0.035 | 0.003 | 0.068 | 0.708 | 0.668 | 0.747 | 0.042 | 0.014 | 0.069 | -0.430 | -0.436 | -0.423 | 0.800 | 0.756 | 0.844 |
| 8 | -0.977 | -0.977 | -0.977 | -0.168 | -0.207 | -0.130 | 0.071 | 0.027 | 0.114 | 0.964 | 0.915 | 1.014 | 0.054 | 0.018 | 0.089 | -0.722 | -0.728 | -0.715 | 1.093 | 1.039 | 1.148 |
| 9 | -0.487 | -0.487 | -0.487 | 0.415 | 0.394 | 0.437 | 0.050 | 0.011 | 0.089 | 0.603 | 0.563 | 0.643 | 0.026 | -0.005 | 0.057 | -0.312 | -0.320 | -0.304 | 0.714 | 0.668 | 0.761 |
| 10 | -0.534 | -0.534 | -0.534 | 0.497 | 0.475 | 0.520 | 0.023 | -0.017 | 0.063 | 0.681 | 0.637 | 0.724 | 0.039 | 0.007 | 0.071 | -0.353 | -0.361 | -0.344 | 0.757 | 0.707 | 0.808 |
| 11 | -0.627 | -0.627 | -0.627 | 0.497 | 0.472 | 0.523 | 0.115 | 0.072 | 0.159 | 0.720 | 0.674 | 0.767 | 0.063 | 0.029 | 0.096 | -0.424 | -0.432 | -0.415 | 0.857 | 0.802 | 0.912 |
| 12 | -0.990 | -0.990 | -0.990 | 0.713 | 0.669 | 0.757 | 0.061 | 0.007 | 0.115 | 0.926 | 0.865 | 0.987 | 0.059 | 0.013 | 0.105 | -0.736 | -0.744 | -0.727 | 1.055 | 0.990 | 1.120 |

**TABLE S7** Bias: PP estimand

| | CCW | | | G-formula | | | MSM | | | Crude ST | | | IPTW ST | | | TD Cox | | | Crude Cox | | |
|---|---|---|---|---|---|---|---|---|---|---|---|---|---|---|---|---|---|---|---|---|---|
| Factor | Est | Lower | Upper | Est | Lower | Upper | Est | Lower | Upper | Est | Lower | Upper | Est | Lower | Upper | Est | Lower | Upper | Est | Lower | Upper |
| 1 | 0.070 | -0.029 | 0.170 | 0.248 | 0.215 | 0.280 | 0.039 | 0.002 | 0.076 | 0.430 | 0.391 | 0.469 | -0.028 | -0.062 | 0.005 | -0.071 | -0.092 | -0.050 | 0.590 | 0.549 | 0.631 |
| 2 | -0.131 | -0.226 | -0.035 | 0.180 | 0.141 | 0.219 | 0.082 | 0.040 | 0.125 | 0.562 | 0.517 | 0.607 | 0.029 | -0.011 | 0.069 | -0.073 | -0.094 | -0.051 | 0.708 | 0.661 | 0.756 |
| 3 | -0.303 | -0.404 | -0.202 | 0.018 | -0.029 | 0.065 | 0.148 | 0.103 | 0.194 | 0.654 | 0.605 | 0.703 | 0.061 | 0.019 | 0.102 | -0.079 | -0.101 | -0.058 | 0.824 | 0.773 | 0.874 |
| 4 | -1.073 | -1.174 | -0.972 | -0.647 | -0.684 | -0.610 | 0.323 | 0.263 | 0.382 | 1.102 | 1.037 | 1.167 | 0.144 | 0.088 | 0.199 | -0.154 | -0.183 | -0.126 | 1.370 | 1.303 | 1.437 |
| 5 | 0.145 | 0.021 | 0.268 | 0.741 | 0.702 | 0.779 | 0.135 | 0.091 | 0.179 | 0.539 | 0.495 | 0.583 | 0.067 | 0.029 | 0.106 | -0.056 | -0.079 | -0.034 | 0.690 | 0.642 | 0.738 |
| 6 | -0.103 | -0.223 | 0.017 | 0.622 | 0.577 | 0.668 | 0.117 | 0.073 | 0.161 | 0.575 | 0.530 | 0.620 | 0.035 | -0.006 | 0.077 | -0.071 | -0.096 | -0.046 | 0.754 | 0.704 | 0.803 |
| 7 | -0.241 | -0.348 | -0.134 | 0.437 | 0.382 | 0.491 | 0.187 | 0.138 | 0.237 | 0.698 | 0.643 | 0.754 | 0.107 | 0.058 | 0.155 | -0.059 | -0.086 | -0.033 | 0.876 | 0.819 | 0.933 |
| 8 | -0.967 | -1.081 | -0.853 | -0.518 | -0.585 | -0.451 | 0.306 | 0.236 | 0.375 | 1.079 | 1.010 | 1.149 | 0.093 | 0.035 | 0.150 | -0.161 | -0.192 | -0.130 | 1.361 | 1.283 | 1.439 |
| 9 | 0.253 | 0.010 | 0.497 | 0.562 | 0.526 | 0.597 | 0.117 | 0.062 | 0.172 | 0.510 | 0.455 | 0.566 | 0.027 | -0.022 | 0.075 | -0.071 | -0.101 | -0.041 | 0.684 | 0.620 | 0.747 |
| 10 | 0.171 | 0.000 | 0.342 | 0.653 | 0.613 | 0.693 | 0.125 | 0.065 | 0.185 | 0.614 | 0.551 | 0.676 | 0.067 | 0.011 | 0.122 | -0.069 | -0.101 | -0.037 | 0.756 | 0.690 | 0.822 |
| 11 | 0.206 | 0.006 | 0.406 | 0.773 | 0.727 | 0.820 | 0.238 | 0.170 | 0.307 | 0.716 | 0.646 | 0.786 | 0.129 | 0.070 | 0.188 | -0.055 | -0.087 | -0.022 | 0.914 | 0.835 | 0.992 |
| 12 | -0.797 | -0.963 | -0.631 | 1.027 | 0.949 | 1.105 | 0.278 | 0.190 | 0.367 | 1.029 | 0.934 | 1.125 | 0.118 | 0.034 | 0.201 | -0.197 | -0.237 | -0.157 | 1.292 | 1.191 | 1.393 |
